# A versatile data repository for GWAS summary statistics-based downstream genomic analysis of human complex traits

**DOI:** 10.1101/2025.10.01.25337099

**Authors:** Peter Sørensen, Palle Duun Rohde

## Abstract

Genome-wide association studies (GWAS) have identified thousands of loci associated with complex traits and diseases, yet the biological interpretation of these findings remains limited. We developed gact, an R package that integrates GWAS summary statistics with diverse genomic resources to facilitate the discovery of causal genes, pathways, and disease mechanisms. The package enables construction of a local database linking variants to genes, biological pathways, protein complexes, and drug–gene interactions, thereby supporting downstream analyses such as fine-mapping, polygenic scoring, and gene set enrichment. Applying gact to large-scale GWAS of coronary artery disease (CAD) and type 2 diabetes (T2D), we identified 142 and 577 significant genes, respectively, including canonical loci for T2D (*PATJ, DEAF1*) and CAD (*OLIG1*), as well as pleiotropic genes such as *TCF7* and *HNF1B*. Bayesian gene set analyses revealed distinct biological signatures—lipid and vascular remodeling pathways in CAD versus β-cell and islet biology in T2D— together with shared enrichment in extracellular matrix and immune signaling. Polygenic score (PGS) analyses demonstrated higher predictive accuracy for CAD than T2D, consistent with differences in common-variant heritability and GWAS power. Partitioned PGS further delineated T2D subgroups through archetypal clustering, separating individuals with predominantly inflammatory versus metabolic risk profiles. These results establish gact as a versatile platform for integrating genomic resources and advancing the biological interpretation of GWAS. By linking genetic associations to biological pathways and subtypes, gact enables a deeper understanding of disease heterogeneity and informs future precision medicine strategies.

## Introduction

Genomic data are forecast to be a key resource for understanding variation in the human phenome and hold great promise for disease diagnostics, prevention, and improved medical intervention. To understand complex human traits and diseases at the molecular genetic level, we need to establish a comprehensive overview of the genomic entities implicated in the manifestation of phenotypic variation and disease susceptibility. Since gene–phenotype links can be derived from many different types of biological experiments and data sources - each growing at an immensely fast rate - there is a need for computational tools that can semi-automatically and efficiently integrate heterogeneous evidence of genomic associations. To this end, we have developed the R package *gact*, designed to establish and populate a comprehensive database focused on genomic associations with complex human traits and diseases.

Genome-wide association studies (GWAS) constitute a valuable tool for understanding the biology of complex human traits and diseases, and thousands of genetic loci have now been identified^1^. Despite these significant advancements, the majority of GWAS loci do not directly implicate the causal genes underlying the associations, which substantially limits the biological insights gained into common disease mechanisms^2,3.^ As a result, many multifaceted efforts have been made to address the challenge of biological interpretability. GWAS summary data - i.e., measures of statistical association between each tested genetic variant and trait variability - are now widely available through public repositories such as the National Human Genome Research Institute (NHGRI)–European Bioinformatics Institute (EBI) GWAS Catalog^4^, GWAS Central^5^, and GWAS Atlas^6^. These resources enable the aggregation of very large GWAS sample sizes through meta-analyses^7^. GWAS summary data can be leveraged to gain insight into the biology of complex traits and diseases by performing fine-mapping^8,9,^ estimating key genetic architecture parameters^10^, computing genome-wide and pathway-specific polygenic scores (PGS)^11,12,^ and conducting gene set enrichment analyses^13^.

Linking genetic variation to biological entities is a nontrivial task, and there are many ways in which genes can be aggregated - such as through gene ontologies^14,15,^ biological pathways^16,17,^ protein complexes^18^, and protein-drug interactions^19,20^ etc. As genetic variation also affects the levels of intermediate molecular phenotypes, establishing these links is crucial for understanding the biology of complex human traits and diseases. One such resource is the Genotype-Tissue Expression (GTEx) project, which provides a comprehensive characterization of the effects of genetic variation on the human transcriptome^21^. While all of these resources are highly valuable, they are not directly integrated, and the information is not stored in a format optimized for efficient downstream statistical genetic analysis— significantly hindering the biological interpretation of genomic associations. With the *gact* package, we have developed a tool that largely circumvents this challenge.

The package serves two primary functions: infrastructure creation and data acquisition. It facilitates the assembly of a structured repository that includes GWAS summary data, all rigorously curated to ensure high data quality. The package integrates a broad spectrum of genomic entities, encompassing genes, proteins, and a variety of biological complexes—such as chemicals and proteins—as well as multiple biological pathways. It is designed to support the biological interpretation of genomic associations, shedding light on their complex relationships in the context of complex trait genomics.

The *gact* package is developed as an R package—the free software environment for statistical computing and graphicsc^22^ —to ensure accessibility for non-specialist users. Its infrastructure is designed to align with that of the qgg package, an R environment for large-scale genomic analysis of complex traits and diseases^23,24.^ In this paper, we describe the gact resource, which aims to enable researchers to discover new connections between genetic entities—such as biological pathways—and complex human traits or diseases. We illustrate the utility of the *gact* package through a genomic analysis of type 2 diabetes and coronary artery disease using GWAS summary data.

## Materials and methods

The current implementation of gact R package creates a local structured database containing gene annotation and gene marker sets based on a range of different biological databases, and aid the user with acquiring new GWAS summary data, performing rigorious quality control of the GWAS data and match the summary data with database structure. The Supplementary File contains details on all statistical genetic models implemented within the package, and how to utilise them.

### Genetic reference data and gene annotation

The user can utilise any genetic reference data of their own choice. The gact package comes with the default setting of utilising genetic reference data from the 1000 Genomes Project^25^. The 1000G data encompass genetic variation across three super-populations; European (EUR), East Asian (EAS), and South Asian (SAS). Initial quality control of the genetic variants was performed with the qgg package^23,24^ such that any genetic variant with a minor allele frequency below 1%, a call rate lower than 95%, and deviation from Hardy-Weinberg proportions (*P*-value<1×10^-12^) was excluded. Furthermore, genetic variations located within the major histocompatibility complex, exhibiting ambiguous alleles (*i.e.*, GC or AT), being multi-allelic, or representing indels, were also excluded^26^. In total 7,235,359 genetic variants passed these quality metrics. The degree of linkage disequilibrium (LD) among pairwise genetic variants were computed in window sizes of 2500 genetic variants.

Genetic variants located within 35kb upstream and 10kb downstream of the open reading frame were linked to that specific gene to include probable regulatory regions^27,28^. In the gact package Ensemble gene annotations were utilised^29^ which can be obtained from: ftp.ensembl.org/pub/grch37/current/gtf/homo_sapiens/Homo_sapiens.GRCh37.87.gtf.gz.

### Mapping genetic variants to gene sets

One of the core features of the gact package is the automatic creation of gene sets; *i.e.*, aggregating genetic variants within annotated genes. In this initial release of the gact package, the following gene sets are automatically generated based on the initial Ensemble gene annotation as described above:

- Protein and transcript gene sets based on the Ensembl database^29^.
- Regulatory genomic feature sets from the Ensembl Regulation database ^29^.
- Gene Ontology (GO) gene sets utilising the GO database ^14,15.^
- Pathway gene sets generated from Reactome^16^ and KEGG databases^17^.
- Protein complex gene sets from the STRING database^18^.
- Small molecule chemical complex gene sets from the STITCH database^30,31.^
- Drug-gene interaction gene sets obtained from the DrugBank database^19,20.^
- Drug ATC (Anatomical Therapeutic Chemica) gene sets based on the ATC^32^ and DrugBank databases^19,20.^
- Gene sets based on drug complexes by combining information from the STRING database^18^ and from DrugBank^19,20.^
- Known disease-gene sets based on experiments, textmining and knowledge base obtained from the DISEASE database^33,34.^

In addition to these gene sets, the gact package also generate gene sets based on known expression quantitative trait loci (eQTLs). The eQTL gene sets were generated from the Genotype-Tissue Expression (GTEx) project^21^ based on version 8 eQTL data downloaded from Google Cloud Storage at: https://storage.googleapis.com/adult-gtex/bulk-qtl/v8/single-tissue-cis-qtl/GTEx_Analysis_v8_eQTL.tar Finally, gene sets based on previous GWAS genome-wide significant genetic variant associations were created based on the GWAS catalog database (data version 1.0 downloaded 2023/04/25, see https://www.ebi.ac.uk/gwas/docs/file-downloads).

### Ingestion of new GWAS summary data

After establishing the database infrastructure, external GWAS summary data can be ingested. Users may download complete GWAS summary statistics, which are then processed by a function that applies user-defined quality control, aligns alleles and SNP effects with genetic reference data, updates the R database object with new information, and records the ingested data in an associated text file. Importantly, if the gact database is established using 1000 Genomes data, the genome build of the entire database structure is GRCh37. Therefore, any GWAS summary data not based on GRCh37 must be lifted to this build. In the accompanying Supplementary Data File, we provide an example illustrating how GWAS summary data can easily be lifted from, for example, GRCh38 to GRCh37.

### Application within UK Biobank

For PGS calculations, we used imputed genetic variants from the UK Biobank (UKB)^35^. We restricted the dataset to unrelated individuals of White British ancestry, excluding participants with more than 5,000 missing genotypes or evidence of autosomal aneuploidy, resulting in 335,532 individuals. Genotypes were hard-called from imputed dosages using a probability threshold of 0.7, and variants with an imputation quality score ≥0.8 were retained using PLINK 2.0^36^. Further quality control excluded variants with minor allele frequency <0.01, call rate <0.95, deviation from Hardy–Weinberg equilibrium (*P* < 1 × 10–12), ambiguous alleles (GC/AT), multi-allelic sites, and indels, while retaining the major histocompatibility complex (MHC) region given its known disease associations. After quality control, 6,627,732 single nucleotide polymorphisms (SNPs) were available for PGS analyses.

Disease case definitions were based on ICD10 codes from the UK Biobank “Diagnosis – main ICD10” data field, supplemented with corresponding self-reported disease codes. Individuals without the relevant ICD10 diagnosis code for a given condition were assigned as controls. For coronary artery disease (CAD), cases were defined using ICD10 codes I21–I25 and self-reported code 1075, yielding a total of 34,726 cases and 300,806 controls. For type 2 diabetes (T2D), cases were defined using ICD10 code E11 together with self-reported codes 1220 and 1223, resulting in 25,828 cases and 309,704 controls.

To explore potential heterogeneity among T2D cases, we performed archetypal analysis in R using the archetypes package (v3.2.1). As input, we constructed individual-level profiles of polygenic scores restricted to the 33 Reactome pathways that showed significant enrichment among T2D cases in the pathway-PGS analyses. Only individuals classified as T2D cases were included in this step. The analysis was carried out on standardized pathway-PGS scores (mean = 0, SD = 1 across cases) to ensure comparability across pathways. Archetypes, representing extreme and interpretable profiles of genetic risk across pathways, were estimated using the alternating least squares (ALS) algorithm implemented in the stepArchetypes function. Models with 2–10 archetypes were fitted, and the optimal number of archetypes was selected based on a combination of residual sum of squares (RSS) inspection and the elbow criterion. Each T2D case was then assigned to the archetype with the highest membership proportion, yielding genetically informed subtypes of T2D.

## Results

### Functionality and Performance of the gact R Package

To evaluate the functionality of the gact R package, we conducted a series of benchmark tests and validation analyses using publicly available GWAS summary statistics and reference data from the 1000 Genomes Project. The package successfully created a structured local database integrating gene annotations and multiple biologically relevant gene sets (Figure 1), as described in the Materials and Methods section.

**Figure 1.**
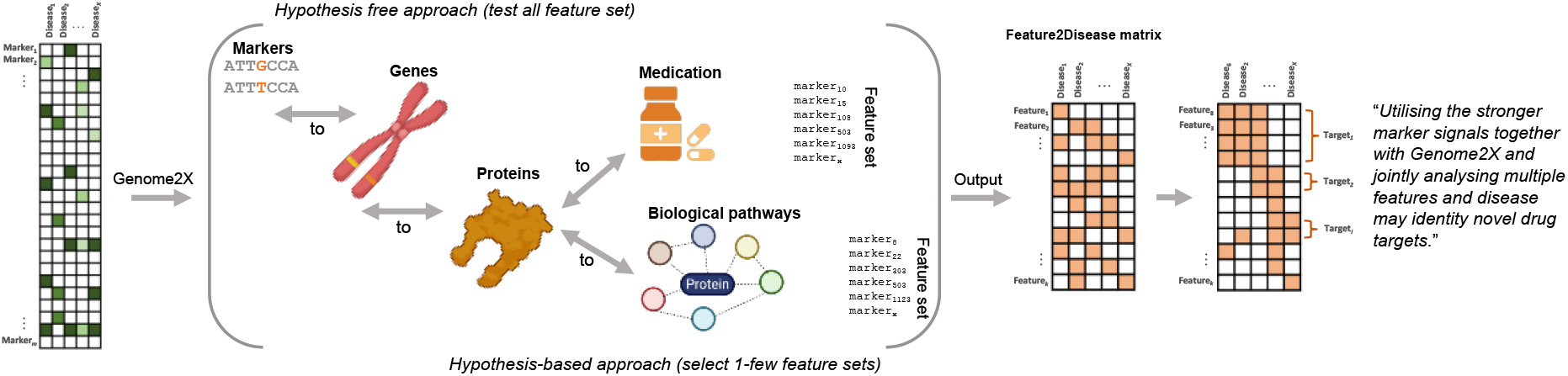
Integrative framework of the gact R package for biological interpretation of genomic associations. The gact workflow supports both hypothesis-free (variant-level) and hypothesis-based (gene set–level) approaches. GWAS-derived genetic variants are mapped to diverse biological entities, including genes, proteins, metabolites, and curated biological pathways. These mappings enable aggregation of association signals into structured gene sets derived from resources such as Reactome, KEGG, STRING, STITCH, and DrugBank. Downstream analyses generate feature/disease matrices that can be used for heritability partitioning, gene-level testing, polygenic scoring, and multi-trait analyses. By leveraging shared genetic architecture across traits and jointly analyzing multiple features and diseases, gact facilitates the identification of disease mechanisms and potential drug targets.

#### Database Construction and Annotation Mapping

The initial database construction completed in approximately 10 minutes on a standard workstation, resulting in a structured repository of approximately 10 million quality-controlled genetic variants mapped to 19,000+ annotated genes. In the current default release, gene set construction incorporated data from 11 external databases, including Ensembl, GO, Reactome, STRING, and DrugBank, yielding over 13 distinct gene sets. The built-in SNP quality control pipeline, based on the qgg package^23,24^, filtered out low-quality variants efficiently (MAF thresholds, call rate, or Hardy-Weinberg equilibrium violations).

#### User Interface and Workflow Efficiency

The package provides a streamlined workflow for users, from data import to gene set generation. Key functions are modular and well-documented, allowing flexible customization. Runtime profiling indicated that the full pipeline—from raw GWAS summary statistics to annotated gene sets—can be completed in under 3 minutes for datasets with up to 10 million variants.

### Application to cardiometabolic traits

To illustrate the utility of the gact package, we applied it to GWAS summary data for coronary artery disease (CAD) and type 2 diabetes (T2D). These traits are well-suited for demonstration because they share genetic architecture yet represent biologically distinct disease processes.

#### Global and Partitioned Heritability

We first estimated SNP-heritability 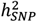 using LDSC. CAD and T2D showed genome-wide heritability estimates of 0.23 (SE=0.03) and 0.32 (SE=.04), respectively. We estimated the genetic correlation (*r*_*g*_) between CAD and T2D to 0.31 (SE=0.06). Partitioning of SNP-heritability by functional annotations demonstrated significant enrichment in promoter-flanking regions and enhancers, with these elements explaining a disproportionately large proportion of the total heritability. This enrichment is consistent with the notion that regulatory elements, particularly those involved in transcriptional regulation, contribute substantially to the genetic architecture of cardiometabolic traits (Supplementary Figure S2)^37^. These results demonstrate the capacity of gact to integrate LD-based heritability models with functionally informed annotations.

#### Gene-Level Associations

Using the VEGAS framework within gact, we identified 142 significant genes for CAD and 577 for T2D after multiple testing correction at *P*_FDR_<0.01 (Supplementary Table S1). A total of 25 genes were significantly associated with both diseases. These included canonical T2D genes such as *PATJ*, and *DEAF1*, reflecting established roles in β-cell biology^38^, and CAD-associated genes such as *OLIG1*, which may represent novel cardiovascular biology. Shared loci such as *TCF7, HNF1B, IFITM3*, and *GPATCH8* highlight pleiotropic pathways linking metabolic and vascular risk, consistent with recent large-scale cross-trait analyses^39,40^. The strongest T2D-specific associations were observed at *PATJ* and *DEAF1*, both of which overlap known islet regulatory processes^41^, while top CAD-specific signals included *OLIG1, GNPTAB*, and *TCEA1*, pointing towards potentially novel mechanisms beyond established lipid biology. The distribution of shared associations along the diagonal (Figure S3) suggests a partially overlapping polygenic basis for T2D and CAD. Importantly, the modular design of gact facilitates integration with external resources such as GTEx, enabling tissue-specific expression annotation to further contextualize gene-level signals and distinguish established from potentially novel loci.

#### Bayesian Gene Set Analysis

We next applied Bayesian MAGMA to evaluate enrichment of curated gene sets, here exemplified with Reactome pathways. We have previously demonstrated the flexibility of this framework by applying BLR-MAGMA to KEGG-derived pathways^42^ and to drug–gene interaction sets^43^, highlighting its utility across diverse biological annotations.

In the multi-trait Bayesian analysis, we identified several gene sets with high posterior inclusion probability (PIP >0.9) in at least one trait (Figure S4). These included pathways reflecting extracellular matrix remodeling (collagen fibril crosslinking, metalloproteinase activation), vascular and endothelial signaling (VEGF interactions, nitric oxide–stimulated guanylate cyclase), and immune-related processes (defensins, antigen presentation, neuroinflammation). Of note, some of the strongest shared signals mapped to general metabolic processes such as glycerophospholipid biosynthesis and cytochrome remodeling, underscoring common mechanisms bridging metabolic dysfunction and vascular disease.

Collectively, these findings reinforce that CAD gene-level associations are largely driven by dyslipidemia and vascular remodeling, while T2D signals cluster around pancreatic islet biology. The multi-trait framework highlights intermediate shared pathways, particularly those linking inflammation, extracellular matrix biology, and metabolic signaling, illustrating how leveraging pleiotropy across related diseases can reveal convergent mechanisms that may be missed in single-trait analyses.

Collectively, these findings reinforce that CAD gene-level associations are largely driven by dyslipidemia and vascular remodeling, while T2D signals cluster around pancreatic islet biology. The multi-trait framework highlights intermediate shared pathways, particularly those linking inflammation, extracellular matrix biology, and metabolic signaling, illustrating how leveraging pleiotropy across related diseases can reveal convergent mechanisms that may be missed in single-trait analyses. Importantly, the Bayesian MAGMA module within gact provides a unified and flexible framework for performing both single- and multi-trait analyses, making it straightforward to uncover disease-specific as well as pleiotropic pathway enrichments.

#### Polygenic Scores

We next evaluated the predictive performance of PGS constructed using the Bayesian linear regression output from fine-mapping. Prediction accuracy was sensitive to both the choice of linkage disequilibrium (LD) reference and variant selection thresholds, with custom LD sets generated in gact performing on par with, and in some cases exceeding, those derived from external resources such as UK Biobank^35^ and Pickrell reference panels^44^ (Figure S7). Across traits, predictive accuracy was higher for CAD compared to T2D, consistent with previous reports of stronger common-variant heritability for CAD (~ 55–60%) vs. T2D (~ 25–30%) in family/twin studies^45^, as well as more extensive variant discovery and greater GWAS sample sizes for CAD allowing more powerful PGS construction^46^. As expected, stricter MAF thresholds (e.g. MAF >0.05) yielded more stable performance, while inclusion of rarer variants provided only marginal gains in model fit.

#### Pathway-Partitioned PGS and Genetic Subtypes

To capture heterogeneity within T2D cases, we constructed pathway-partitioned PGS based on Reactome pathways. Several pathways showed significant PGS enrichment in T2D patients compared to controls, including those with well-established relevance for T2D (Figure 2A). Notably, the *Signaling by TCF7L2 mutants* pathway emerged as enriched, consistent with the pivotal role of *TCF7L2*, one of the strongest and most consistently replicated T2D risk loci, in β-cell function and incretin signaling^47^. We also observed enrichment of *Transcriptional regulation by VENTX*, which involves regulators such as *IL6, p53*, and *CDKN2A*, linking genetic associations to processes of inflammation and cellular senescence – mechanisms increasingly implicated in insulin resistance and β-cell failure^48^. In addition, enrichment was observed for *Tyrosine catabolism*, in line with metabolomics studies that report altered aromatic amino acid metabolism as a precursor of diabetes onset^49^.

**Figure 2.**
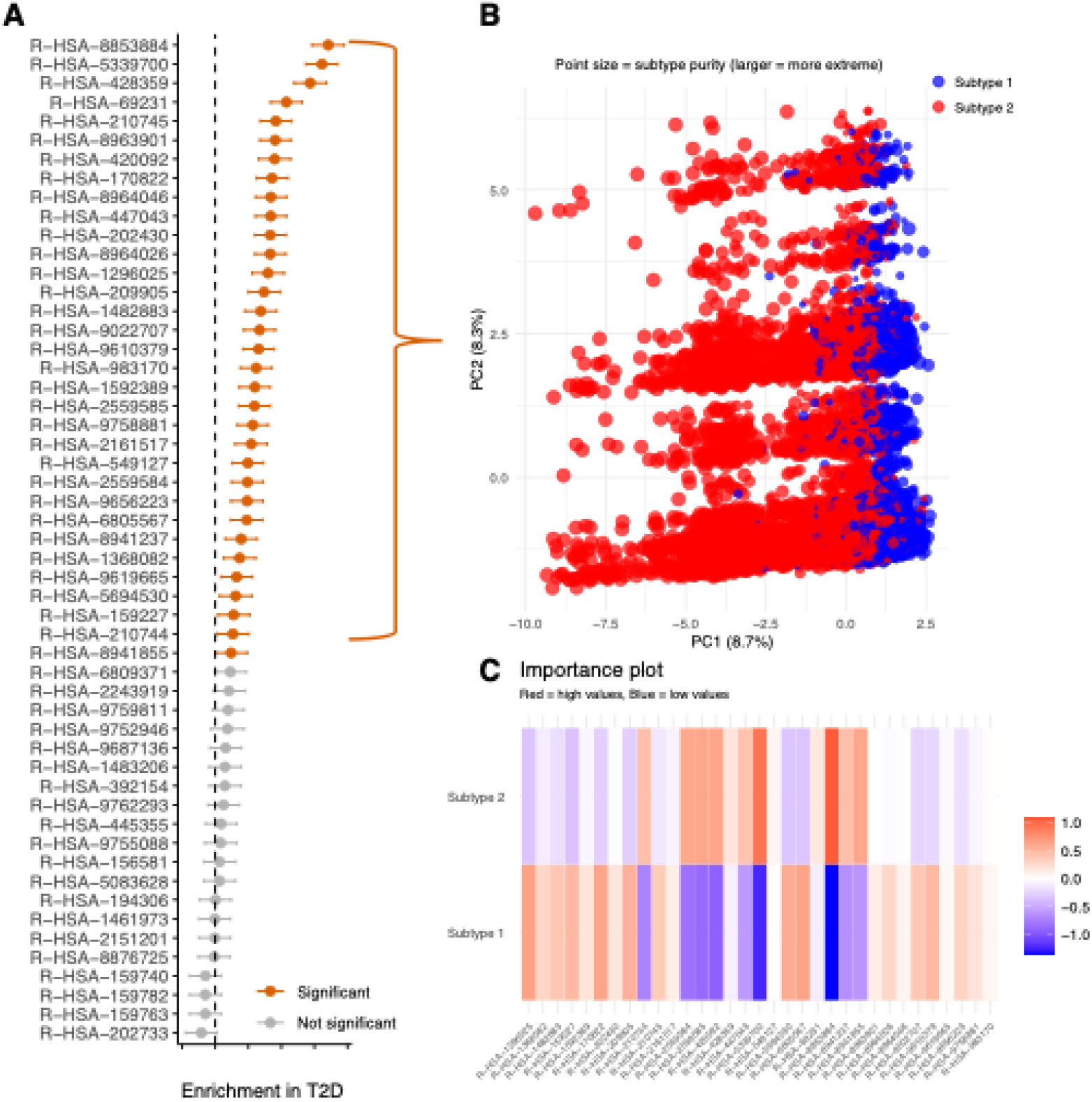
Identification of genetically defined subtypes in type 2 diabetes using pathway-partitioned polygenic scores. (**A**) Enrichment of genetic burden across Reactome pathways in T2D patients compared to controls; significantly enriched pathways are highlighted in orange. (**B**) Genetic subtypes of T2D cases identified via archetypal analysis of the enriched pathways; Subtype 1 (blue) and Subtype 2 (red) are shown, with point size reflecting subtype purity. (**C**) Importance plot illustrating the contribution of each pathway to the definition of the genetic subtypes; red indicates high contribution and blue low contribution.

To further dissect the heterogeneity of genetic risk, we applied archetypal analysis to the pathway-level PGS derived from Reactome gene sets significantly enriched in T2D. This unsupervised approach identified two major archetypes of T2D cases (Figure 2B). Visualization of cases in principal component space revealed that individuals segregated into two partially overlapping but distinct clusters, with point size reflecting subtype purity (Figure 2B). Subtype 1 was characterized by lower scores across immune- and inflammation-related pathways, whereas Subtype 2 displayed elevated genetic risk for pathways linked to inflammatory signaling, cellular stress responses, and metabolism (Figure 2C).

The separation of cases into these archetypes highlights that genetic risk for T2D is not uniformly distributed but instead aggregates into biologically coherent subgroups. Importantly, the distinct contribution of inflammatory versus metabolic pathways mirrors current understanding of T2D as a multifactorial disease with both β-cell dysfunction and systemic insulin resistance components.

## Discussion

In this study, we present the gact R package as a flexible and extensible framework for building structured genomic databases and performing downstream statistical genetic analyses. By integrating variant-level quality control, gene annotation, and a broad collection of gene set resources, gact enables researchers to move seamlessly from raw GWAS summary statistics to biologically contextualized results.

Application of gact to cardiometabolic traits highlights its ability to recapitulate known disease biology while also pointing to novel insights. The gene-level and pathway-based analyses confirmed canonical associations for T2D (e.g., PATJ, DEAF1, TCF7L2) and CAD (e.g., OLIG1, GNPTAB), while uncovering pleiotropic signals linking inflammation, extracellular matrix remodeling, and lipid metabolism across both traits. These results underscore the utility of multi-trait Bayesian gene set models implemented in gact for identifying convergent mechanisms that might be overlooked in single-trait analyses.

The pathway-partitioned polygenic score framework further illustrates the added value of gact in dissecting disease heterogeneity. In T2D, enrichment of pathways related to incretin signaling, inflammation, and amino acid metabolism not only validated established biology but also enabled stratification of patients into genetically distinct subgroups. This type of analysis provides a promising avenue for precision medicine, where genetic risk can be translated into biologically informed disease subtypes.

While the current implementation of gact is already comprehensive, several limitations merit consideration. First, the reliance on a fixed reference genome build (GRCh37) necessitates liftover of newer GWAS data, which can introduce uncertainty. Second, although multiple functional and pathway databases are integrated, their coverage and accuracy are inherently limited by the underlying curation. Future updates will aim to incorporate additional reference panels (e.g., TOPMed, gnomAD) and to extend support for more recent genome builds.

In conclusion, gact provides a unified computational environment for genomic data integration, quality control, and statistical analysis. By combining flexible database construction with advanced tools for gene-level, pathway-level, and polygenic risk analyses, gact offers a platform well suited to studying the genetic architecture of complex diseases. Its application to cardiometabolic traits illustrates the package’s potential to reveal both disease-specific and shared biological pathways, and to move toward genetically informed disease stratification.

## Data availability

All summary-level results generated in this study are provided in the Supplementary Material. The individual-level genotype and phenotype data used in this work were obtained from the UK Biobank (application ID 96479) and are available to bona fide researchers upon application to the UK Biobank (https://www.ukbiobank.ac.uk/).

## Acknowledgments

This research has been conducted using the UK Biobank Resource under application number 96479.

## References

1. Visscher, P.M., Wray, N.R., Zhang, Q., Sklar, P., McCarthy, M.I., Brown, M.A., and Yang, J. (2017). 10 Years of GWAS Discovery: Biology, Function, and Translation. Am. J. Hum. Genet. 101, 5–22. 10.1016/j.ajhg.2017.06.005.

2. Donnelly, P. (2008). Progress and challenges in genome-wide association studies in humans. Nature 456, 728–731. 10.1038/nature07631.

3. Gallagher, M.D., and Chen-Plotkin, A.S. (2018). The Post-GWAS Era: From Association to Function. Am. J. Hum. Genet. 102, 717–730. 10.1016/j.ajhg.2018.04.002.

4. Sollis, E., Mosaku, A., Abid, A., Buniello, A., Cerezo, M., Gil, L., Groza, T., Güneş, O., Hall, P., Hayhurst, J., et al. (2022). The NHGRI-EBI GWAS Catalog: knowledgebase and deposition resource. Nucleic Acids Res. 51, D977–D985. 10.1093/nar/gkac1010.

5. Beck, T., Rowlands, T., Shorter, T., and Brookes, A.J. (2022). GWAS Central: an expanding resource for finding and visualising genotype and phenotype data from genome-wide association studies. Nucleic Acids Res. 51, D986–D993. 10.1093/nar/gkac1017.

6. Liu, X., Tian, D., Li, C., Tang, B., Wang, Z., Zhang, R., Pan, Y., Wang, Y., Zou, D., Zhang, Z., et al. (2022). GWAS Atlas: an updated knowledgebase integrating more curated associations in plants and animals. Nucleic Acids Res. 51, gkac924.. 10.1093/nar/gkac924.

7. Yengo, L., Vedantam, S., Marouli, E., Sidorenko, J., Bartell, E., Sakaue, S., Graff, M., Eliasen, A.U., Jiang, Y., Raghavan, S., et al. (2022). A saturated map of common genetic variants associated with human height. Nature 610, 704–712. 10.1038/s41586-022-05275-y.

8. Stephens, M., and Balding, D.J. (2009). Bayesian statistical methods for genetic association studies. Nat. Rev. Genet. 10, 681–690. 10.1038/nrg2615.

9. Faye, L.L., Machiela, M.J., Kraft, P., Bull, S.B., and Sun, L. (2013). Re-Ranking Sequencing Variants in the Post-GWAS Era for Accurate Causal Variant Identification. PLoS Genet. 9, e1003609. 10.1371/journal.pgen.1003609.

10. Moser, G., Lee, S.H., Hayes, B.J., Goddard, M.E., Wray, N.R., and Visscher, P.M. (2015). Simultaneous discovery, estimation and prediction analysis of complex traits using a Bayesian mixture model. PLoS Genet. 11, e1004969. 10.1371/journal.pgen.1004969.

11. Lloyd-Jones, L.R., Zeng, J., Sidorenko, J., Yengo, L., Moser, G., Kemper, K.E., Wang, H., Zheng, Z., Magi, R., Esko, T., et al. (2019). Improved polygenic prediction by Bayesian multiple regression on summary statistics. Nat. Commun. 10, 5086. 10.1038/s41467-019-12653-0.

12. Choi, S.W., García-González, J., Ruan, Y., Wu, H.M., Porras, C., Johnson, J., Consortium, B.D.W. group of the P.G., Hoggart, C.J., and O’Reilly, P.F. (2023). PRSet: Pathway-based polygenic risk score analyses and software. PLOS Genet. 19, e1010624. 10.1371/journal.pgen.1010624.

13. Leeuw, C.A. de, Neale, B.M., Heskes, T., and Posthuma, D. (2016). The statistical properties of gene-set analysis. Nat. Rev. Genet. 17, 353–364. 10.1038/nrg.2016.29.

14. Ashburner, M., Ball, C.A., Blake, J.A., Botstein, D., Butler, H., Cherry, J.M., Davis, A.P., Dolinski, K., Dwight, S.S., Eppig, J.T., et al. (2000). Gene Ontology: tool for the unification of biology. Nat. Genet. 25, 25–29. 10.1038/75556.

15. Aleksander, S.A., Balhoff, J., Carbon, S., Cherry, J.M., Drabkin, H.J., Ebert, D., Feuermann, M., Gaudet, P., Harris, N.L., Hill, D.P., et al. (2023). The Gene Ontology knowledgebase in 2023. Genetics 224, iyad031. 10.1093/genetics/iyad031.

16. Milacic, M., Beavers, D., Conley, P., Gong, C., Gillespie, M., Griss, J., Haw, R., Jassal, B., Matthews, L., May, B., et al. (2023). The Reactome pathway knowledgebase 2024. Nucleic Acids Res. 52, D672–D678. 10.1093/nar/gkad1025.

17. Kanehisa, M., and Goto, S. (2000). KEGG: Kyoto Encyclopedia of Genes and Genomes. Nucleic Acids Res. 28, 27–30. 10.1093/nar/28.1.27.

18. Szklarczyk, D., Gable, A.L., Lyon, D., Junge, A., Wyder, S., Huerta-Cepas, J., Simonovic, M., Doncheva, N.T., Morris, J.H., Bork, P., et al. (2019). STRING v11: protein–protein association networks with increased coverage, supporting functional discovery in genome-wide experimental datasets. Nucleic Acids Res. 47, D607–D613. 10.1093/nar/gky1131.

19. Knox, C., Wilson, M., Klinger, C.M., Franklin, M., Oler, E., Wilson, A., Pon, A., Cox, J., Chin, N.E. (Lucy), Strawbridge, S.A., et al. (2023). DrugBank 6.0: the DrugBank Knowledgebase for 2024. Nucleic Acids Res. 52, D1265–D1275. 10.1093/nar/gkad976.

20. Wishart, D.S., Knox, C., Guo, A.C., Shrivastava, S., Hassanali, M., Stothard, P., Chang, Z., and Woolsey, J. (2006). DrugBank: a comprehensive resource for in silico drug discovery and exploration. Nucleic Acids Res. 34, D668–D672. 10.1093/nar/gkj067.

21. Consortium, T. Gte., Aguet, F., Anand, S., Ardlie, K.G., Gabriel, S., Getz, G.A., Graubert, A., Hadley, K., Handsaker, R.E., Huang, K.H., et al. (2020). The GTEx Consortium atlas of genetic regulatory effects across human tissues. Science 369, 1318–1330. 10.1126/science.aaz1776.

22. Team, R.C. (2023). R: A language and environment for statistical ## computing.

23. Rohde, P.D., Sørensen, I.F., and Sørensen, P. (2019). qgg: an R package for large-scale quantitative genetic analyses. Bioinformatics 36, 2614–2615. 10.1093/bioinformatics/btz955.

24. Rohde, P.D., Sørensen, I.F., and Sørensen, P. (2023). Expanded utility of the R package, qgg, with applications within genomic medicine. Bioinformatics 39, btad656. 10.1093/bioinformatics/btad656.

25. Auton, A., Abecasis, G.R., Altshuler, D.M., Durbin, R.M., Abecasis, G.R., Bentley, D.R., Chakravarti, A., Clark, A.G., Donnelly, P., Eichler, E.E., et al. (2015). A global reference for human genetic variation. Nature 526, 68–74. 10.1038/nature15393.

26. Marees, A.T., Kluiver, H. de, Stringer, S., Vorspan, F., Curis, E., Marie-Claire, C., and Derks, E.M. (2018). A tutorial on conducting genome-wide association studies: Quality control and statistical analysis. Int. J. Methods Psychiatr. Res. 27, e1608. 10.1002/mpr.1608.

27. Maston, G.A., Evans, S.K., and Green, M.R. (2006). Transcriptional Regulatory Elements in the Human Genome. Annual Review of Genomics and Human Genetics 7, 29–59. 10.1146/annurev.genom.7.080505.115623.

28. Trubetskoy, V., Pardiñas, A.F., Qi, T., Panagiotaropoulou, G., Awasthi, S., Bigdeli, T.B., Bryois, J., Chen, C.-Y., Dennison, C.A., Hall, L.S., et al. (2022). Mapping genomic loci implicates genes and synaptic biology in schizophrenia. Nature 604, 502–508. 10.1038/s41586-022-04434-5.

29. Martin, F.J., Amode, M.R., Aneja, A., Austine-Orimoloye, O., Azov, A.G., Barnes, I., Becker, A., Bennett, R., Berry, A., Bhai, J., et al. (2022). Ensembl 2023. Nucleic acids Res. 51, D933–D941. 10.1093/nar/gkac958.

30. Kuhn, M., Mering, C. von, Campillos, M., Jensen, L.J., and Bork, P. (2008). STITCH: interaction networks of chemicals and proteins. Nucleic Acids Res. 36, D684–D688. 10.1093/nar/gkm795.

31. Szklarczyk, D., Santos, A., von Mering, C., Jensen, L.J., Bork, P., and Kuhn, M. (2016). STITCH 5: augmenting protein–chemical interaction networks with tissue and affinity data. Nucleic Acids Res. 44, D380–D384. 10.1093/nar/gkv1277.

32. Methodology, W.C.C. for D.S. (2024). ATC classification index with DDDs. Oslo, Norway.

33. Pletscher-Frankild, S., Pallejà, A., Tsafou, K., Binder, J.X., and Jensen, L.J. (2015). DISEASES: Text mining and data integration of disease–gene associations. Methods 74, 83–89. 10.1016/j.ymeth.2014.11.020.

34. Grissa, D., Junge, A., Oprea, T.I., and Jensen, L.J. (2022). Diseases 2.0: a weekly updated database of disease–gene associations from text mining and data integration. Database: J. Biol. Databases Curation 2022, baac019. 10.1093/database/baac019.

35. Bycroft, C., Freeman, C., Petkova, D., Band, G., Elliott, L.T., Sharp, K., Motyer, A., Vukcevic, D., Delaneau, O., O’Connell, J., et al. (2018). The UK Biobank resource with deep phenotyping and genomic data. Nature 562, 203–209. 10.1038/s41586-018-0579-z.

36. Chang, C.C., Chow, C.C., Tellier, L.C., Vattikuti, S., Purcell, S.M., and Lee, J.J. (2015). Second-generation PLINK: rising to the challenge of larger and richer datasets. GigaScience 4, 7. 10.1186/s13742-015-0047-8.

37. Bulik-Sullivan, B.K., Loh, P.-R., Finucane, H.K., Ripke, S., Yang, J., Patterson, N., Daly, M.J., Price, A.L., and Neale, B.M. (2015). LD Score regression distinguishes confounding from polygenicity in genome-wide association studies. Nat. Genet. 47, 291–295. 10.1038/ng.3211.

38. Thurner, M., Bunt, M. van de, Torres, J.M., Mahajan, A., Nylander, V., Bennett, A.J., Gaulton, K.J., Barrett, A., Burrows, C., Bell, C.G., et al. (2018). Integration of human pancreatic islet genomic data refines regulatory mechanisms at Type 2 Diabetes susceptibility loci. eLife 7, e31977. 10.7554/elife.31977.

39. Kurt, Z., Cheng, J., Barrere-Cain, R., McQuillen, C.N., Saleem, Z., Hsu, N., Jiang, N., Pan, C., Franzén, O., Koplev, S., et al. (2023). Shared and distinct pathways and networks genetically linked to coronary artery disease between human and mouse. eLife 12, RP88266. 10.7554/elife.88266.

40. Hageh, C.A., O’Sullivan, S., Platt, D.E., Henschel, A., Chacar, S., Gauguier, D., Abchee, A., Alefishat, E., Nader, M., and Zalloua, P.A. (2024). Coronary artery disease patients with rs7904519 (TCF7L2) are at a persistent risk of type 2 diabetes. Diabetes Res. Clin. Pr. 207, 111052. 10.1016/j.diabres.2023.111052.

41. Varshney, A., Scott, L.J., Welch, R.P., Erdos, M.R., Chines, P.S., Narisu, N., Albanus, R.D., Orchard, P., Wolford, B.N., Kursawe, R., et al. (2017). Genetic regulatory signatures underlying islet gene expression and type 2 diabetes. Proc. Natl. Acad. Sci. 114, 2301–2306. 10.1073/pnas.1621192114.

42. Gholipourshahraki, T., Bai, Z., Shrestha, M., Hjelholt, A., Hu, S., Kjolby, M., Rohde, P.D., and Sørensen, P. (2024). Evaluation of Bayesian Linear Regression models for gene set prioritization in complex diseases. PLOS Genet. 20, e1011463. 10.1371/journal.pgen.1011463.

43. Hjelholt, A.J., Gholipourshahraki, T., Bai, Z., Shrestha, M., Kjølby, M., Sørensen, P., and Rohde, P.D. (2025). Leveraging Genetic Correlations to Prioritize Drug Groups for Repurposing in Type 2 Diabetes. medRxiv, 2025.06.13.25329590. 10.1101/2025.06.13.25329590.

44. Berisa, T., and Pickrell, J.K. (2015). Approximately independent linkage disequilibrium blocks in human populations. Bioinformatics 32, 283–285. 10.1093/bioinformatics/btv546.

45. Ehret, G.B. (2013). Framingham’s Contribution to Gene Identification for CV Risk Factors and Coronary Disease. Glob. Hear. 8, 59–65. 10.1016/j.gheart.2012.12.010.

46. Grace, C., Hopewell, J.C., Watkins, H., Farrall, M., and Goel, A. (2022). Robust estimates of heritable coronary disease risk in individuals with type 2 diabetes. Genet. Epidemiology 46, 51–62. 10.1002/gepi.22434.

47. Bosque-Plata, L. del, Martínez-Martínez, E., Espinoza-Camacho, M.Á., and Gragnoli, C. (2021). The Role of TCF7L2 in Type 2 Diabetes. Diabetes 70, 1220–1228. 10.2337/db20-0573.

48. Bowker, N., Shah, R.L., Sharp, S.J., Luan, J., Stewart, I.D., Wheeler, E., Ferreira, M.A.R., Baras, A., Wareham, N.J., Langenberg, C., et al. (2020). Meta-analysis investigating the role of interleukin-6 mediated inflammation in type 2 diabetes. EBioMedicine 61, 103062. 10.1016/j.ebiom.2020.103062.

49. Floegel, A., Stefan, N., Yu, Z., Mühlenbruch, K., Drogan, D., Joost, H.-G., Fritsche, A., Häring, H.-U., Angelis, M.H. de, Peters, A., et al. (2013). Identification of Serum Metabolites Associated With Risk of Type 2 Diabetes Using a Targeted Metabolomic Approach. Diabetes 62, 639–648. 10.2337/db12-0495.

