## Supplementary File for "A versatile data repository for GWAS summary statistics-based downstream genomic analysis of human complex traits"

#### General workflow of the gact package

**8 Maintaining and updating the gact package** **23**

**Citered literature** **25**

### 1 Introduction

This document is a supplementary companion to the scientific paper describing the `gact` (**G**enomic **A**ssociations of **C**omplex **T**raits) package, providing additional details, examples, and usage instructions.

In this document, we will cover how to:

- Initialise the `gact` database
- Populate the database with new GWAS summary statistics

Following this, we will demonstrate the main types of genomic analyses implemented in `gact` by analysing type 2 diabetes (T2D) and coronary artery disease (CAD) using publicly available GWAS summary data. This includes:

- Global and partitioned estimates of heritability ($\hat{h}^2_{SNP}$) and genetic correlations ($\hat{r}_g$)
- Gene-level associations using the VEGAS-approach
- Gene-set enrichment analyses
- Statistical fine mapping of causal genetic variants
- Polygenic scores and their partitioning across biological-enriched gene sets

### 2 The gact database

We highly recommend users to create an `R`-environment using mamba, as shown below, to ensure that all dependencies and package version are as required by the `gact`- and `qgg`-package (Rohde et al., 2019, 2023).

```
conda install -n base -c conda-forge mamba

mamba create -n rgact \
  r-base \
  r-devtools \
  r-remotes \
  r-xml \
  r-rcurl \
  r-pkgbuild \
  zlib \
  compilers \
  make \
  pkg-config \
  libblas=*=*mkl \
  -c conda-forge
```

At the time of writing this document, this installs

```
> sessionInfo()
  R version 4.4.3 (2025-02-28)
  Platform: x86_64-conda-linux-gnu
```

```
Running under: AlmaLinux 9.4 (Seafoam Ocelot)
```

##### 2.1 Initialise the database

If the gact database has not been established this is the first step (see our GitHub for more details gact package). This only has to be done, as more prior data and GWAS summary data easily can be inserted into the database.

```
library(gact)
  GAlist <- gact(version="hsa.0.0.1", dbdir="../gact", task="download")
  saveRDS(GAlist, file=".../gact/hsa.0.0.1/GAlist_hsa.0.0.1.rds")
```

All subsequent data management steps and analyses requires genetic reference data. Here we utilise the 1000G (1kG) data as our backbone. To use 1kG the following must be done:

```
# Download 1000G data (if not allready downloaded)
  GAlist <- downloadDB(GAlist=GAlist, what="1000G")
  saveRDS(GAlist, file=".../gact/hsa.0.0.1/GAlist_hsa.0.0.1.rds")
```

To be able to use the 1kG data, we first need to create a qgg Glist-object (please consult our GitHub page for the qgg package for details).

```
#Prepare Glist for 1000G data for European ancestry (EUR)

# Marker IDs in database
  rsids <- GAlist$rsids

# Define the file paths for the original bed/bim/fam files to read
  bedfiles <- file.path(GAlist$dirs["marker"], "g1000_eur.bed")
  bimfiles <- file.path(GAlist$dirs["marker"], "g1000_eur.bim")
  famfiles <- file.path(GAlist$dirs["marker"], "g1000_eur.fam")

# Define the file paths for the filtered bed/bim/fam files to write
  bedfiles_filtered <- file.path(GAlist$dirs["marker"], "g1000_eur_filtered.bed")
  bimfiles_filtered <- file.path(GAlist$dirs["marker"], "g1000_eur_filtered.bim")
  famfiles_filtered <- file.path(GAlist$dirs["marker"], "g1000_eur_filtered.fam")

# Call the writeBED function to filter and write the data
  writeBED(bedRead=bedfiles,
           bimRead=bimfiles,
           famRead=famfiles,
           bedWrite=bedfiles_filtered,
           bimWrite=bimfiles_filtered,
           famWrite=famfiles_filtered,
           rsids=rsids)
```

```
# Prepare summary (i.e. Glist) for 1000G genotype data
  Glist <- gprep(study="1000G EUR",
                 bedfiles=bedfiles_filtered,
                 bimfiles=bimfiles_filtered,
                 famfiles=famfiles_filtered)

# Save Glist for use later
  saveRDS(Glist, file=file.path(GAlist$dirs["marker"],"Glist_1000G_eur_filtered.rds"))
```

The final step before ingesting GWAS summary data into the database is to compute sparse linkage disequilibrium (LD) and LD scores. This is a computational demanding step when the number of SNPs and samples become very large.

```
# Load GAlist
  GAlist <- readRDS(file=".../gact/hsa.0.0.1/GAlist_hsa.0.0.1.rds")

# Load Glist with information about genotypes in 1000G
  Glist <- readRDS(file=file.path(GAlist$dirs["marker"],"Glist_1000G_eur_filtered.rds"))

# Marker IDs used in sparse LD computation
  rsids <- unlist(Glist$rsids)

# Compute Sparse LD matrix and LD scores for EAS and save for later use
  Glist <- gprep(Glist, task = "sparseld", msize = 1000, rsids = rsids, overwrite = FALSE)

  saveRDS(Glist, file=file.path(GAlist$dirs["marker"],"Glist_1000G_eur_filtered.rds"))

  markers <- data.frame(rsids=unlist(Glist$rsids),
                        chr=unlist(Glist$chr),
                        pos=unlist(Glist$pos),
                        ea=unlist(Glist$a1),
                        nea=unlist(Glist$a2),
                        eaf=unlist(Glist$af),
                        maf=unlist(Glist$maf),
                        map=unlist(Glist$map),
                        ldscores=unlist(Glist$ldscores))
  rownames(markers) <- markers$rsids
  fwrite(markers, file=file.path(GAlist$dirs["marker"],"markers_1000G_eur_filtered.txt.gz"))
```

##### 2.2 Ingesting GWAS summary data into the database

When the `gact`-database has been established as shown above, it is time to start populating/ingested GWAS summary data into the database. This task is done using the `updateStatDB()`, which input all necessary data into the database. Before running `updateStatDB()` the `stat`-object should be created which contains the GWAS summary data. Below we'll showcase how this can be done for the two example traits.

```
GAlist <- updateStatDB(GAlist = GAlist,
                       stat = stat,                  # R-object
                       source = "...",               # markers file name
                       trait = "...",                # name of trait
                       type = "...",                 # quantitative or binary
                       gender = "...",               # male, female, or both
                       ancestry = "...",             # ancestry
                       build = "...",                # genome build, GRCh37 or GRCh38
                       reference = "...",            # PMID
                       n = NA,                       # total sample size
                       ncase = NA,                   # if binary, how many cases
                       ncontrol = NA,                # if binary, how many controls
                       comments = "..."              # comments
                       )
```

##### 2.3 Example: Ingesting CAD and T2D GWAS summary data

For T2D we are using the GWAS summary data from Mahajan 2018 unadjusted for BMI and without UK Biobank subjects (as this allow us to use the GWAS summary data later for constructing PGS within UKB) (Mahajan et al., 2018). The T2D GWAS data was downloaded from Diagram consortium (link). Importantly, the GWAS summary data must be in GRCH37 genome build (as this is the genome build of 1kG), and the order and names of the columns in the `stat`-object should be as shown below. A key feature of the `updateStatDB`-function is, that it aligns alleles and their effects according to the reference genetic data within the database.

```
# Load GWAS data
  fname_stat <- "./Mahajan.NatGenet2018b.T2D-noUKBB.European.zip"
  stat <- fread(fname_stat, data.table = FALSE)

# Modify columns according to required format
# Subset and rename columns according to required format
  stat <- stat[, c("SNP", "Chr", "Pos", "EA", "NEA", "Beta", "SE", "Pvalue")]
  colnames(stat) <- c("marker", "chr", "pos", "ea", "nea", "b", "seb", "p")

# Update database
  GAlist <- updateStatDB(GAlist = GAlist,
                         stat = stat,
                         source = "Mahajan.NatGenet2018b.T2D-noUKBB.European.zip",
                         trait = "T2D",
                         type = "binary",
                         gender = "both",
                         ancestry = "EUR",
                         build = "GRCh37",
                         reference = "PMID:30297969",
                         n = 456236,
                         ncase = 55927,
                         ncontrol = 400309,
                         comments = "Exclude UK biobank",
```

```
                          writeStatDB = TRUE)

# Save updated database
  saveRDS(GAlist, file = "./gact/hsa.0.0.1/GAlist_hsa.0.0.1.rds", compress = FALSE)
```

Beside the Mahajan et al. (2018) T2D GWAS, we also utilise the Cardiogram CAD GWAS (link). The database contains a text-file that list all the GWAS summary data that has been ingested within the database, as shown in Table S1.

**Table S1:** Example of the overview file generated by `gact`. For each GWAS/trait included in the database, the table reports phenotype name, abbreviation, reference, year, sample size (N), and case-control breakdown where relevant, ancestry, genome build and possible user-defined notes.

| id | trait | type | gender | ncase | ncontrol | neff | reference | ancestry | build | comments |
|---|---|---|---|---|---|---|---|---|---|---|
| GWAS1 | T2DM | binary | both | 55927 | 400309 | 49071.27 | PMID:30297969 | EUR | GRCh37 | Exclude UKB |
| GWAS2 | CAD | binary | both | 60801 | 123504 | 40743.15 | PMID:26343387 | EUR | GRCh37 | Exclude UKB |

### 3 Estimating genetic parameters

#### 3.1 Estimation from GWAS summary data

LD score regression (LDSC) is a statistical technique used to estimate the heritability and genetic correlation among complex traits using GWAS summary statistics (Bulik-Sullivan et al., 2015). It examines the relationship between LD scores — which reflect how strongly single nucleotide polymorphisms (SNPs) are correlated with neighbouring genetic variants — and GWAS summary statistics. This method distinguishes genuine polygenic signals from confounding factors such as population stratification, thereby enabling the quantification of the contribution of common variants to trait heritability (see Box 3.1).

**Box 3.1: LDSC**

LDSC is based on the observation that under a polygenic model, SNPs that are in high LD with many other SNPs, tend to capture more heritability. So, their test statistics (e.g., $\chi^2$) tend to be inflated — not necessarily because they are causal, but because they 'tag' more causal variants.

The expected GWAS test statistic for SNP $j$ is modelled as:

$$\mathbb{E}[\chi_j^2] = 1 + \frac{Nh^2\ell_j}{M} + a \quad (1)$$

where:

- $\chi_j^2$ is the chi-squared statistic for SNP $j$,
- $N$ is the GWAS sample size,
- $h^2$ is the SNP-heritability (heritability explained by all SNPs),
- $\ell_j = \sum_k r_{jk}^2$ is the LD score of SNP j (sum of squared correlations with nearby SNPs),
- $M$ is the number of SNPs analysed,

- $a$ is the intercept, accounting for confounding effects (e.g., population stratification, cryptic relatedness).

LDSC regresses the observed $\chi^2$ values on LD scores $\ell_j$. The slope of this regression estimates $\frac{Nh^2}{M}$, from which $h^2$ can be obtained. The intercept, $a$ estimates the average contribution of confounding to test statistic inflation.

LDSC can also estimate the genetic correlation $r_g$ between two traits using a bivariate extension. For SNP $j$, the expected product of $Z$-scores from two GWAS is:

$$\mathbb{E}[Z_{1j}Z_{2j}] = \frac{r_g\sqrt{N_1h_1^2N_2h_2^2} \cdot \ell_j}{M} + a_{12} \quad (2)$$

where:

- $Z_{1j}$ and $Z_{2j}$ are $Z$-scores for SNP $j$ in traits 1 and 2,
- $h_1^2$, $h_2^2$ are the SNP heritabilities for traits 1 and 2,
- $r_g$ is the genetic correlation,
- $N_1$, $N_2$ are the respective sample sizes,
- $\ell_j$ is the LD score,
- $M$ is the number of SNPs,
- $a_{12}$ is the intercept that captures shared confounding (e.g., sample overlap).

The slope of the regression of $Z_{1j}Z_{2j}$ on $\ell_j$ is proportional to the genetic correlation $r_g$.

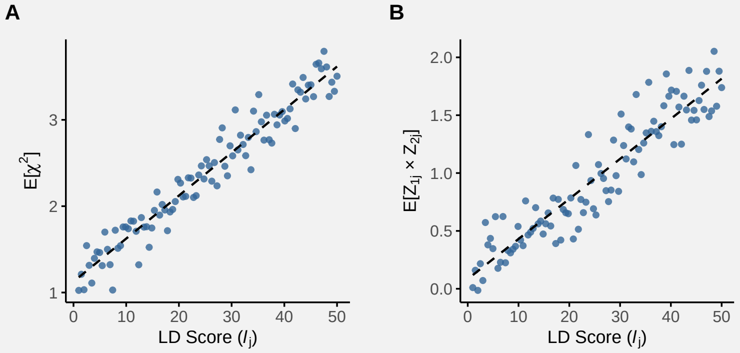

**Figure: LDSC regression for estimating heritability and genetic correlation**. (**A**) In univariate LD score regression, chi-squared statistics from a GWAS are regressed on LD Scores to estimate SNP heritability. The slope reflects the contribution of polygenic signal, while the intercept captures inflation due to confounding. (**B**) In bivariate LD score regression, the product of Z-scores from two GWAS is regressed on LD Scores to estimate genetic correlation. The slope corresponds to the genetic covariance, standardized by the heritabilities of each trait.

LDSC yields valid estimates of SNP heritability and genetic correlation under several key assumptions:

1. **Polygenic Architecture**
   - The trait is influenced by many SNPs with small effects (i.e., infinitesimal model).
   - Causal variants are randomly distributed across the genome (or their distribution is

independent of LD score).

2. **No Confounding Between LD Score and Bias**
   - Confounding factors (e.g., population stratification, cryptic relatedness, or batch effects) affect all SNPs equally, regardless of LD Score.
   - This ensures the intercept of the LDSC regression captures such confounding, while the slope captures true heritability.
3. **LD Scores are Accurate**
   - LD Scores are precomputed from a reference panel (like 1000 Genomes).
   - The reference population's LD structure must match the study population's ancestry.
4. **No Sample Overlap (for Genetic Correlation)**
   - When estimating genetic correlation between two traits, independent samples are preferred.
   - Sample overlap leads to inflation in the covariance term unless properly modelled.
5. **Independence Between LD and Effect Size**
   - There should be no systematic relationship between LD score and the magnitude of effect sizes (i.e., SNPs in high-LD regions are not systematically more likely to be causal).

##### 3.2 Example: Estimating genetic parameters for CAD and T2D

Estimating genetic parameters as SNP-based heritability and genetic correlation between two traits is easily obtained with the `gact`-package as population-specific LD scores ($\ell_j$) is present within the database when the sparse LD-matrix has been computed.

```
# Load GAlist
  GAlist <- readRDS(file=".../gact/hsa.0.0.1/GAlist_hsa.0.0.1.rds")

# Select GWAS study IDs
  studyIDs <- c("GWAS1","GWAS2")

# Get GWAS summary statistics for studyIDs (e.g. z and n) from gact database
  stat <- getMarkerStat(GAlist=GAlist, studyID=studyIDs)

# Get ldscores matched to the ancestry of GWAS data
  ldscores <- getLDscoresDB(GAlist=GAlist, ancestry="EUR", version="1000G")

# Estimate heritability and genetic correlation using using ldsc
# and estimate standard error (SE) with jackknife bootstrap
  fit.h2 <- ldsc(z=stat$z, n=stat$n, ldscores=ldscores, what="h2", SE.h2=T)
  fit.rg <- ldsc(z=stat$z, n=stat$n, ldscores=ldscores, what="rg", SE.rg=T)
```

For CAD and T2D, the SNP-based heritability estimates are shown in Figure S1A, and the estimated genetic correlation between the two traits is shown in Figure S1B.

In addition, we implemented a Bayesian version of standard LDSC. This allows for a flexible framework to obtain partitioned estimates of SNP-based heritability ($\hat{h}^2_{\mathrm{SNP}_{\mathrm{set}}}$). By using the `ldsc` function with `method = "bayesC"`, heritability estimation is performed using a BayesC prior within the BLR framework.

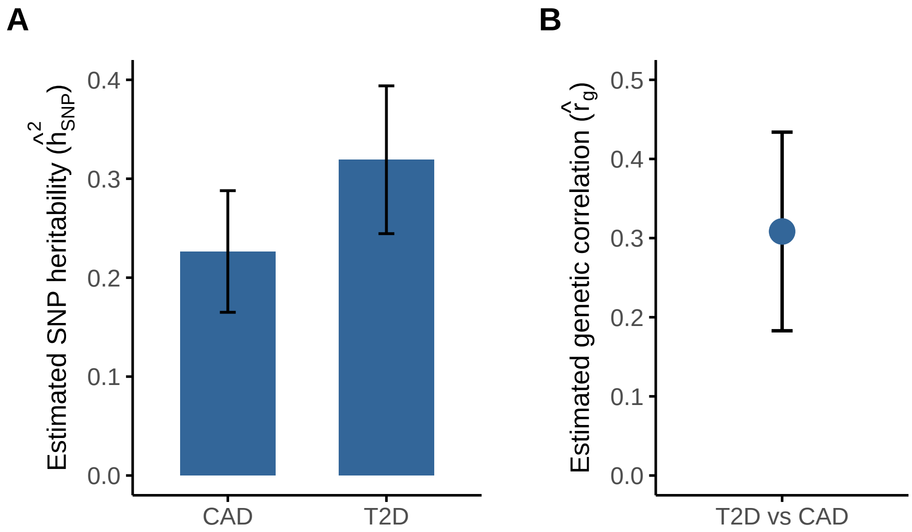

**Figure S1: Estimated SNP heritability and genetic correlation from LDSC**. (**A**) SNP heritability ($\hat{h}^2_{\mathrm{SNP}}$) for coronary artery disease (CAD) and type 2 diabetes (T2D), with 95% confidence intervals. (**B**) Genetic correlation ($\hat{r}_g$) between CAD and T2D, estimated using bivariate LDSC. Error bars indicate 95% confidence intervals.

An integral component of the `gact` package is its streamlined approach to linking genetic variants with diverse genomic features. For example, the command `getMarkerSets(GAlist=GAlist, feature="Regulatory Categories")` maps genetic variants in the database to regulatory categories. These sets can then be used in `ldsc` to estimate the proportion of phenotypic variance attributable to variants within predefined functional categories. Note that not all variants will be assigned to a feature. Therefore, we recommend using the argument `residual = TRUE`, which generates an additional marker set containing all variants not included in any predefined category.

```
# Partitioned h2 across chromosomes
  sets <- getMarkerSets(GAlist=GAlist, feature="Chromosomes")
  fit <- ldsc(z=stat$z, n=stat$n, ldscores=ldscores, sets=sets, what="h2",
              method="bayesC", residual=TRUE)

# Partitioned h2 across regulatory categories
  sets <- getMarkerSets(GAlist=GAlist, feature="Regulatory Categories")
  fit <- ldsc(z=stat$z, n=stat$n, ldscores=ldscores, sets=sets, what="h2",
              method="bayesC", residual=TRUE)
```

Figure S2 shows the partitioned SNP-based heritability ($\hat{h}^2_{\mathrm{SNP}_{\mathrm{set}}}$) estimates across chromosomes (panel **A**) and regulatory categories (panel **B**) for CAD and T2D. As expected, heritability is distributed across all chromosomes, with some variation in contribution size, likely reflecting differences in gene density, LD structure, and trait architecture. Partitioning by regulatory annotations highlights that variants located in promoter-flanking regions and enhancers explain a disproportion-

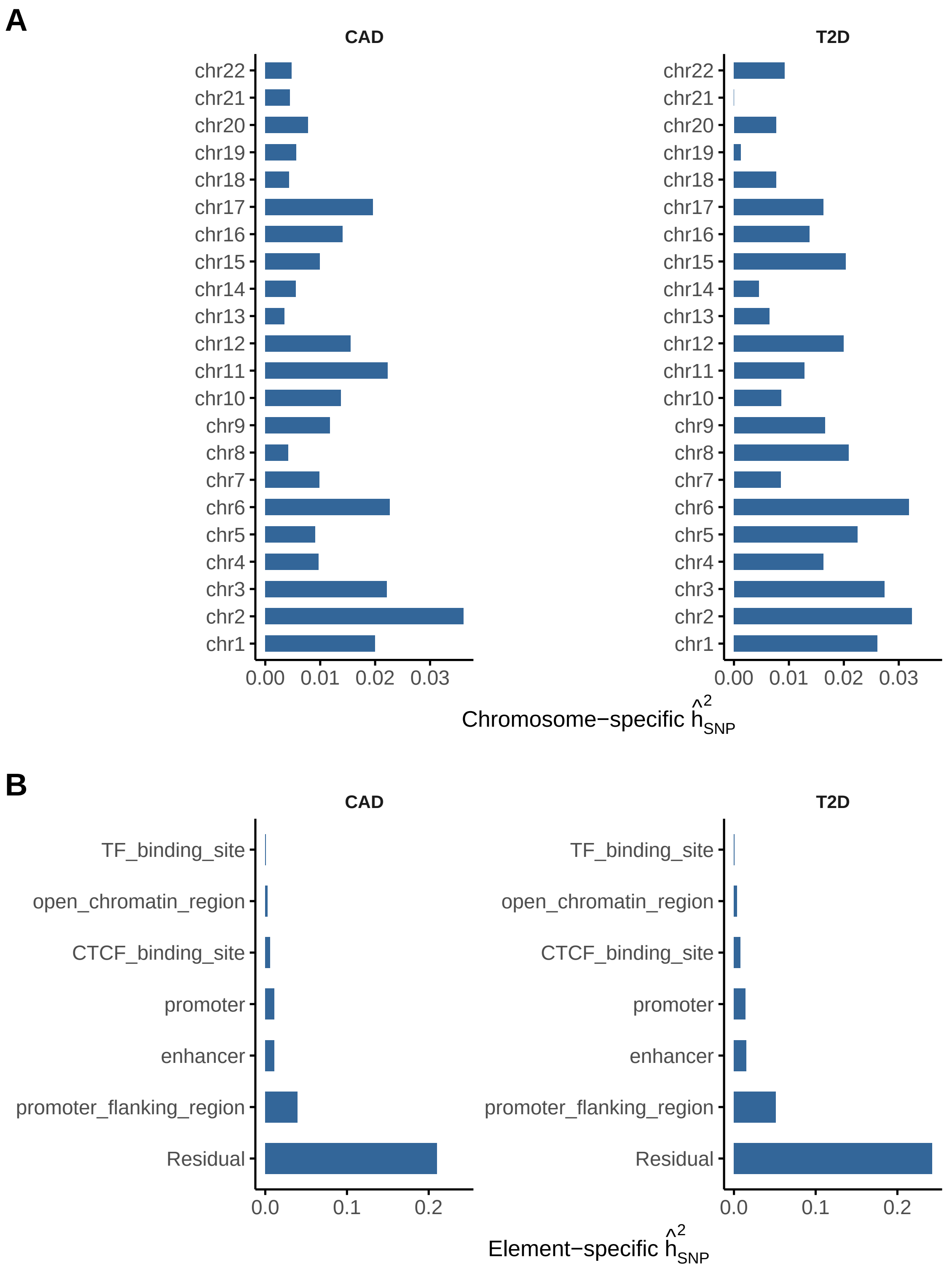

**Figure S2: Partitioned SNP heritability across genomic features for CAD and T2D.** (**A**) Chromosome-specific SNP heritability estimates for coronary artery disease (CAD) and type 2 diabetes (T2D). Heritability estimates are based on stratified LD score regression and reflect the proportion of total SNP-based heritability attributed to each chromosome. (**B**) SNP heritability attributed to selected regulatory element annotations, including promoters, enhancers, and transcription factor binding sites. Estimates are shown separately for CAD and T2D.

ately large share of the total heritability. These findings are consistent with the hypothesis that regulatory elements play a key role in the genetic architecture of cardiometabolic traits.

### 4 Gene-level association

#### 4.1 From associated genetic variants to gene level associations

In GWAS, individual SNPs may not reach genome-wide significance due to small effect sizes or incomplete tagging. However, aggregating evidence across SNPs within a gene can reveal associations at the gene level that would otherwise be missed. But SNPs are not independent; LD induces correlation. Simply summing test statistics (e.g., $\chi^2$-square values) would inflate the type I error rate if correlations are ignored. VEGAS (Versatile Gene-based Association Study) combines SNP-level test statistics within a gene, accounting for LD via the covariance of the Z-statistics (Liu et al., 2010; Mishra & Macgregor, 2015). The gene-level test statistic is a quadratic form in correlated standard normals. While early versions relied on simulations, modern implementations use saddlepoint (see Box 4.1) approximations for computational efficiency and accuracy, especially for small $P$-values.

**Box 4.1: Saddlepoint Estimation**

The term comes from saddlepoint integration in complex analysis, where one evaluates an integral by expanding the exponent of a function around a *saddlepoint*: a point where the first derivative is zero, but the second derivative is not.

In statistics, the saddlepoint is a value $\hat{\xi}$ that satisfies a particular equation involving the cumulant-generating function (CGF), which is a key part of the approximation. Note, the moment-generating function (MGF) of a random variable X is:

$$M_X(\xi) = \mathbb{E}[e^{\xi X}] \tag{3}$$

The cumulant-generating function is simply the logarithm of the MGF:

$$K_X(\xi) = \log \mathbb{E}[e^{\xi X}] = \log M_X(\xi) \tag{4}$$

This is defined for values of $\xi$ such that the expectation exists. The CGF has a very useful property: its derivatives at zero give the cumulants of the distribution.
For example:

- $K'_X(0)$ = mean of $X$
- $K''_X(0)$ = variance of $X$
- $K^{(3)}_X(0)$ = third cumulant (related to skewness)
- $K^{(4)}_X(0)$ = fourth cumulant (related to kurtosis)

Saddlepoint approximation is especially useful when:

- The statistic of interest is a nonlinear function of many random variables (e.g., quadratic forms like sums of correlated chi-squares).
- You need accurate $P$-values in the extreme tail of a distribution (where simulation or standard approximations like normal or chi-squared fail).

- Computational efficiency matters (e.g., evaluating $P$-values for thousands of genes in a GWAS).

Suppose you have a statistic $T$, and you want to compute its right-tail probability:

$$P(T \geq t) \tag{5}$$

If $T$ has a CGF $K(\xi)$, then the saddlepoint $\hat{\xi}$ is the solution to:

$$K\text{'}(\hat{\xi}) = t \tag{6}$$

Then define:

$$w = \text{sign}(\hat{\xi})\sqrt{2(\hat{\xi}t - K(\hat{\xi}))}u = \hat{\xi}\sqrt{K\text{''}(\hat{\xi})} \tag{7}$$

The saddlepoint approximation to the one-sided tail probability is:

$$P(T \geq t) \approx 1 - \Phi(w) + \phi(w)\left(\frac{1}{w} - \frac{1}{u}\right) \tag{8}$$

Where $\Phi(w)$ is the cumulative distribution function (CDF) of the standard normal, and $\phi(w)$ is the probability density function (PDF) of the standard normal.

In VEGAS, the gene-level test statistic is a sum of correlated $\chi^2$ statistics, each corresponding to SNP-level $Z$-statistics within a gene region. Specifically, the gene-level statistic is:

$$T_G = \sum_{j=1}^{m} Z_j^2 \tag{9}$$

where $Z_j \sim \mathcal{N}(0, 1)$ under the null, and the vector $\mathbf{Z} = (Z_1, Z_2, \dots, Z_m)^\top$ follows a multivariate normal distribution with covariance matrix $\Sigma$, reflecting LD among SNPs in the gene.

The null distribution of $T_G$ is not simply $\chi^2$, because of the correlation among SNPs (i.e., LD). The original VEGAS approach simulated this null distribution by drawing thousands of multivariate normal vectors $\mathbf{Z}^{(b)} \sim \mathcal{N}(0, \Sigma)$ and computing:

$$T_G^{(b)} = \sum_{j=1}^{m} \left(Z_j^{(b)}\right)^2 \tag{10}$$

This is computationally expensive, especially for many genes or very low $P$-values. To address this, the recent update of VEGAS uses the saddlepoint approximation (SPA) to obtain accurate $P$-values analytically, avoiding costly simulations.

Let's denote the observed statistic as $T_G = t$. We aim to compute:

$$p = P(T_G \geq t) \tag{11}$$

Since $T_G$ is a quadratic form in normal variables with correlation $\Sigma$, its distribution is a weighted sum of $\chi^2$ variables. Denote the eigenvalues $\lambda_1, \dots, \lambda_m$ of $\Sigma$, then:

$$T_G \overset{d}{=} \sum_{j=1}^{m} \lambda_j \chi_1^2(j) \tag{12}$$

The cumulant generating function (CGF) of $T_G$:

$$K(\xi) = -\frac{1}{2} \sum_{j=1}^{m} \log(1 - 2\xi\lambda_j) \tag{13}$$

This is defined for $\xi < \frac{1}{2\lambda_{\max}}$, where $\lambda_{\max}$ is the largest eigenvalue.

The SPA approximation of $P$-value:

Define:

- $\hat{\xi}$: the saddlepoint, solution to $K$'$(\hat{\xi}) = t$
- $w = \text{sign}(\hat{\xi})\sqrt{2(\hat{\xi}t - K(\hat{\xi}))}$
- $u = \hat{\xi}\sqrt{K\text{''}(\hat{\xi})}$

Then, the saddlepoint-approximated one-sided $P$-value is:

$$P(T_G \geq t) \approx 1 - \Phi(w) + \phi(w)\left(\frac{1}{w} - \frac{1}{u}\right) \tag{14}$$

where:

- $\Phi(\cdot)$ is the standard normal CDF,
- $\phi(\cdot)$ is the standard normal PDF.

This gives an accurate and fast $P$-value even for small $P$-values and avoids the need for simulation.

##### 4.2 Example: Genes associated with T2D and CAD

We performed gene-level association analyses for T2D and CAD using the VEGAS method (with saddlepoint approximation), which aggregates SNP-level association signals while accounting for LD within genes. This approach enables identification of genes that harbour multiple modest-effect variants contributing to disease risk, complementing traditional single-variant analyses.

```
# Load GAlist and Glist with information on 1000G matched to the ancestry of GWAS data
  GAlist <- readRDS(file="./gact/hsa.0.0.1/GAlist_hsa.0.0.1.rds")
  Glist <- readRDS(file.path(GAlist$dirs["marker"],"Glist_1000G_eur_filtered.rds"))

# Extract gene-marker sets (include markers 40kb/10kb upstream/downstream)
  markerSets <- getMarkerSets(GAlist = GAlist, feature = "Genesplus")
```

```
# Select study1
  studyID <- "GWAS1"

# Get GWAS summary statistics from gact database
  stat <- getMarkerStat(GAlist=GAlist, studyID=studyID)

# Check and align summary statistics based on marker information in Glist
  stat <- checkStat(Glist=Glist, stat=stat)

# Gene analysis using VEGAS
  res <- vegas(Glist=Glist, sets=markerSets, stat=stat, verbose=TRUE)
  filename <- file.path(GAlist$dirs["gsea"], paste0(studyID, "_vegas.rds"))
  saveRDS(res,file=filename)
```

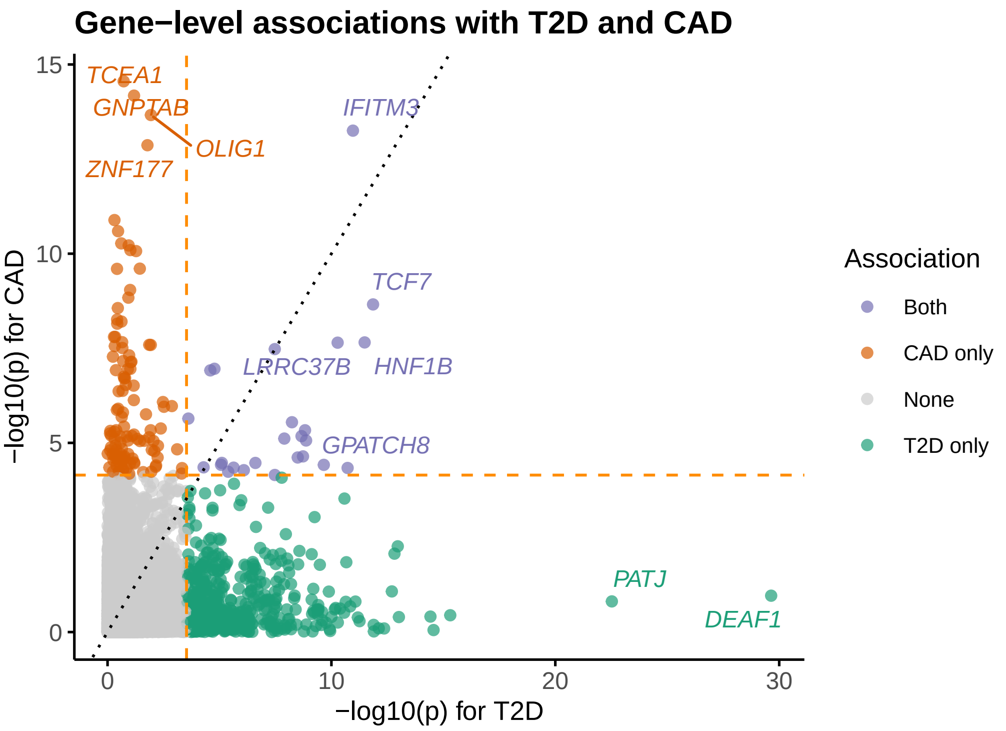

**Figure S3: Gene-level associations for T2D and CAD**. Genes (points) are coloured by their significant association with CAD, T2D or both.

Our results highlight several genes exhibiting significant associations with T2D and CAD (Figure S3), including both shared and trait-specific loci, thereby providing insights into the underlying genetic architecture and potential biology involved in cardiometabolic disease etiology.

### 5 Gene set enrichment analyses

#### 5.1 From genes to gene sets

To further interpret gene-level associations and uncover the biological pathways underlying complex traits, gene set enrichment analysis is a widely used strategy. In the `gact` package, we have implemented a Bayesian version of MAGMA, extending our recent methodological work based on Bayesian Linear Regression (BLR) models (Gholipourshahraki et al., 2024). This approach tests whether predefined gene sets, such as biological pathways, functional annotations, or tissue-specific gene expression profiles, are enriched for genetic association signals. Importantly, it accounts for differences in gene size, LD structure, and uncertainty in gene-level effect estimates, providing a more robust and interpretable framework for enrichment analysis.

Traditional enrichment methods often rely on binary thresholds (e.g., selecting the top $n$ significant genes), potentially discarding valuable information. In contrast, BLR-MAGMA adopts a Bayesian framework that models the full posterior distribution of gene-level effects, allowing more nuanced inference and potentially improved power, particularly for highly polygenic traits.

**Box 5.1: The BLR-MAGMA Model**

The foundation of the model is a linear regression framework that relates per-gene association statistics to gene set membership. Let $\mathbf{y} \in \mathbb{R}^n$ denote the vector of gene-level statistics (e.g., $Z$-scores), where $n$ is the number of genes. The linear model is expressed as:

$$\mathbf{y} = \mathbf{X}\beta + \varepsilon \tag{15}$$

where,

- $\mathbf{X} \in \{0,1\}^{n \times m}$ is a binary design matrix indicating whether gene $i$ belongs to gene set $j$
- $\beta \in \mathbb{R}^m$ is the vector of gene set effects
- $\varepsilon \sim \mathcal{N}(0, \sigma^2 \mathbf{I})$ is the residual noise

To regularize this model, we assume a BayesC prior on the gene set effects:

$$\beta_j \sim (1-\pi)\,\delta_0 + \pi\,\mathcal{N}(0, \sigma_\beta^2) \tag{16}$$

where $\delta_0$ is a point mass at zero and $\pi$ controls the expected proportion of non-zero effects ($\pi = 0.001$ as default). The variance parameter $\sigma_\beta^2$ follows an inverse-chi-squared prior. This spike-and-slab prior enables automatic sparsity and uncertainty quantification over gene set inclusion.

For multi-trait analysis, the model is extended as:

$$\begin{bmatrix}\beta_1 \\ \beta_2\end{bmatrix} = \left( \begin{bmatrix}\mathbf{X}_1^\top \mathbf{X}_1 & 0 \\ 0 & \mathbf{X}_2^\top \mathbf{X}_2\end{bmatrix} + \mathbf{I} \otimes \mathbf{V}_\beta^{-1} \right)^{-1} \begin{bmatrix}\mathbf{X}_1^\top \mathbf{y}_1 \\ \mathbf{X}_2^\top \mathbf{y}_2\end{bmatrix} \tag{17}$$

Here, $\mathbf{V}\beta$ and $\mathbf{V}\varepsilon$ are covariance matrices for the gene set effects and residuals, respectively. These matrices allow for shared effects across traits and account for correlated errors, offering increased power in identifying gene sets with pleiotropic effects.

$$\mathbf{V}\beta = \begin{bmatrix}\sigma^2\beta_1 & \sigma_{\beta_{12}} \\ \sigma_{\beta_{21}} & \sigma_{\beta_2}^2\end{bmatrix}, \quad \mathbf{V}\varepsilon = \begin{bmatrix}\sigma^2\varepsilon_1 & \sigma_{\varepsilon_{12}} \\ \sigma_{\varepsilon_{21}} & \sigma_{\varepsilon_2}^2\end{bmatrix} \tag{18}$$

Both $\mathbf{V}\beta$ and $\mathbf{V}\varepsilon$ follow inverse-Wishart priors, allowing for adaptive regularization and borrowing of information across traits.

The BLR model is estimated using Markov Chain Monte Carlo (MCMC) via the 'blr()' function from the 'qgg' package. For both single- and multi-trait models, we used 3,000 iterations with a 500-iteration burn-in period, and confirmed convergence across multiple chains. Our implementation supports both fixed and user-defined gene sets, and allows joint modeling of multiple traits, making it well-suited for uncovering shared pathways across related cardiometabolic diseases.

##### 5.2 Example: Shared reactome pathways associated with T2D and CAD

To demonstrate the utility of our Bayesian gene set enrichment approach, we applied BLR-MAGMA to a joint analysis of T2D and CAD. These cardiometabolic diseases are known to share genetic architecture and overlapping biological mechanisms. Using gene-level association statistics derived from VEGAS, we tested for enrichment across curated gene sets from the Reactome database—representing well-defined biological pathways and cellular processes. By leveraging the multi-trait BLR model, we account for genetic correlation between traits and enable shared signal detection across traits while preserving trait-specific resolution (see Figure S4 ). This joint modelling increases power to identify pathways involved in pleiotropic effects and may highlight key biological functions underpinning both disorders.

#### 6 Statistical fine mapping

While genome-wide association studies (GWAS), gene-level association testing, and gene set enrichment analyses provide valuable insights into the biological mechanisms underlying complex traits, they do not typically pinpoint the specific causal variants driving these associations. This is a key limitation, as the strongest association signals often arise from non-causal variants in LD with the true causal ones.

Statistical fine mapping aims to resolve this uncertainty by estimating the probability that each genetic variant is causal, given the observed association signals and the LD structure among variants. This process helps to prioritize a smaller subset of variants - often referred to as a credible set - that are most likely to have functional effects on the trait. By moving from broader association signals to likely causal variants, fine mapping provides a critical step toward functional validation and therapeutic targeting.

In the context of our analysis, fine mapping complements the earlier steps by refining the signals observed at the gene and pathway levels. While gene-based tests aggregate association signals across a gene region, and pathway analyses evaluate enrichment across predefined gene sets, fine mapping helps identify the precise variants that may be responsible for driving these signals. This layered approach—from genome-wide to variant-level resolution—supports a more interpretable and actionable understanding of the genetic architecture of complex diseases such as T2D and CAD.

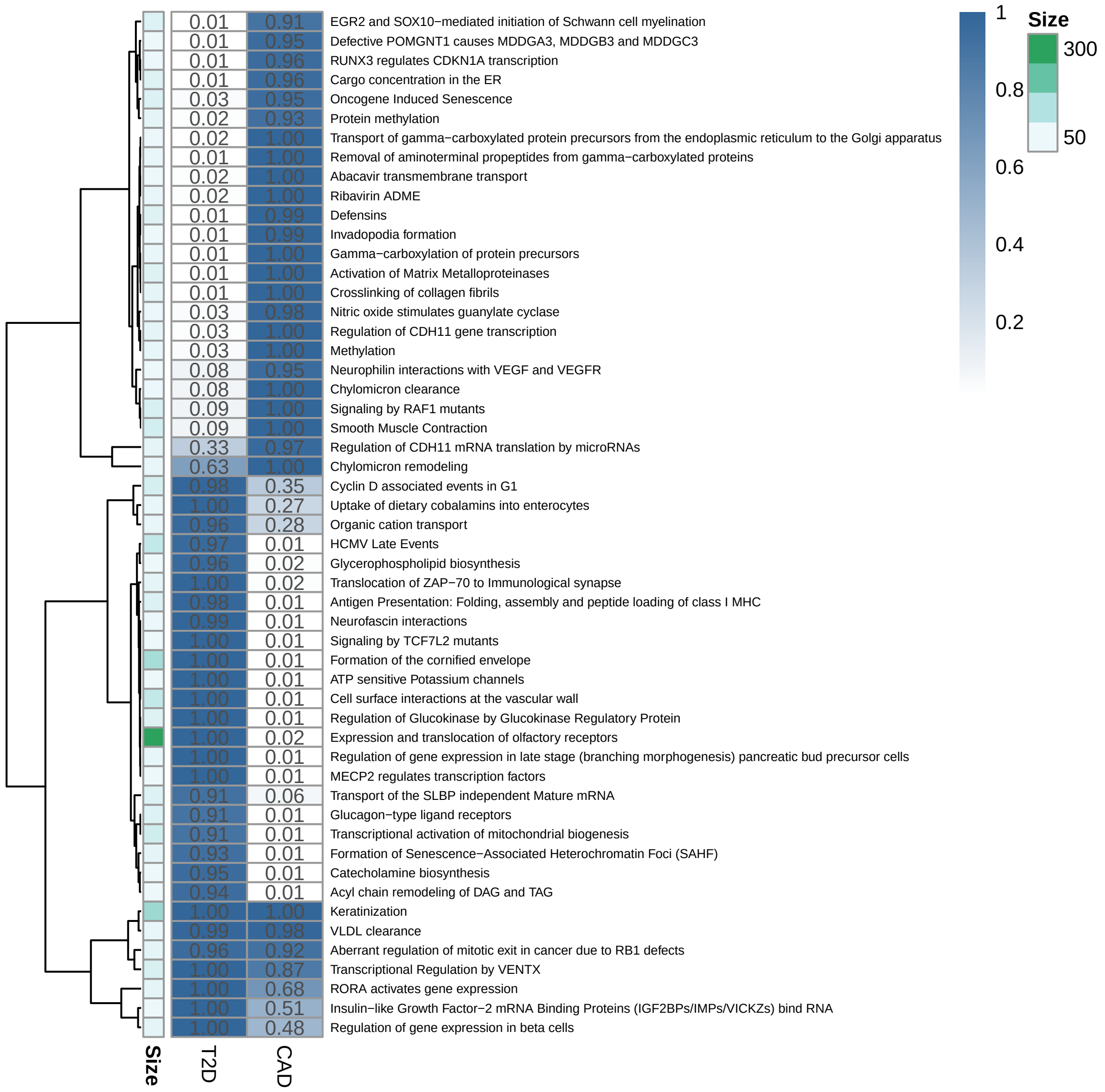

**Figure S4: Top Reactome gene sets identified by multi-trait BLR-MAGMA analysis of T2D and CAD**. The heatmap shows the posterior inclusion probability (PIP) for each gene set–trait pair. Only gene sets with PIP > 0.9 in at least one trait are shown. Color intensity reflects PIP, while numeric labels indicate exact values. Rows are sorted by maximum absolute effect size across traits. An annotation bar indicates the number of genes per pathway.

#### 6.1 Bayesian linear regression models for fine mapping

To improve resolution in identifying causal variants, we employed a statistical fine-mapping framework based on Bayesian Linear Regression (BLR) models, as described in our recent work (Shrestha et al., 2025). BLR models provide a flexible and principled way to jointly model the effects of multiple genetic variants within a locus while accounting for LD and polygenicity. This stands in contrast to traditional single-SNP analyses, which often yield inflated signals due to modelled LD structure.

The core of the BLR approach is a multiple regression model:

$$\mathbf{y} = \mathbf{X}\beta + \epsilon \tag{19}$$

where $\mathbf{y}$ is a vector of phenotypic values (or estimated SNP effects from GWAS summary statistics), $\mathbf{X}$ is the genotype matrix, $\beta$ represents the vector of SNP effect sizes, and $\epsilon \sim \mathcal{N}(0, \sigma^2 I)$ denotes the residual errors.

To enable variable selection and shrinkage, we use the BayesC prior for SNP effects:

$$\beta_j \sim \begin{cases} 0 & \text{with probability } 1 - \pi \\ \mathcal{N}(0, \sigma_\beta^2) & \text{with probability } \pi \end{cases} \tag{20}$$

This spike-and-slab prior assumes that only a small proportion $\pi$ of variants have non-zero effects, allowing the model to distinguish between likely causal and non-causal variants. The prior on $\sigma_\beta^2$ is typically an inverse chi-squared distribution, and inference is performed using Markov Chain Monte Carlo (MCMC) sampling.

An important output of this framework is the Posterior Inclusion Probability (PIP) for each variant, defined as the proportion of MCMC iterations in which the variant is included in the model. PIPs provide a direct probabilistic measure of the likelihood that a variant is causal, facilitating the construction of credible sets—minimal subsets of variants that together account for a predefined proportion (e.g., 95%) of the posterior probability.

Compared to existing fine-mapping approaches, BLR models offer several advantages:

- Joint modelling of all variants in a region, which improves resolution in high LD regions.
- Robust handling of multiple causal variants within a locus.
- Flexible priors that can be tailored to different assumptions about genetic architecture.
- Computational scalability via the `gact` package, which integrates summary statistics and reference LD panels.

Overall, this Bayesian fine-mapping approach provides a powerful tool for prioritizing candidate variants for downstream functional studies and integrates seamlessly with earlier gene- and pathway-level analyses.

Fine mapping is particular sensitive to mismatch in LD between LD reference panel and the GWAS summary statistic LD (please see Shrestha et al. (2025) for discussion on this). In `gact` we handle this by using MCMC eigen value decomposition, which performs robust fine mapping even on summary statistics from GWAS meta analyses where the LD is often different than the reference

LD used. This can be specified by `algorithm="mcmc-eigen"` which we highly recommend users to use.

##### 6.2 Example: Fine mapping of T2D and CAD loci

Having established trait-relevant genes and biological pathways through gene-level association and enrichment analyses, we next sought to pinpoint the specific genetic variants that may drive these associations. To this end, we applied our Bayesian fine-mapping framework to the T2D and CAD summary statistics. Fine mapping enables the identification of credible sets of variants that are most likely to have a causal impact on disease risk, helping to disentangle true signals from correlated non-causal variants in regions of high linkage disequilibrium. By estimating posterior inclusion probabilities (PIPs) for each variant, our approach provides a probabilistic ranking of candidate causal variants and refines the search space for downstream experimental validation. This step is essential to bridge statistical association with biological mechanism and therapeutic target discovery.

For this analysis, we estimated LD using genotypes from the White-British, unrelated subset of the UK Biobank. To ensure computational efficiency and reduce redundancy due to LD, we partitioned the genome into approximately independent LD blocks using a custom algorithm. Specifically, we developed a function (`createLDsets`) that identifies local minima in smoothed LD scores (computed using a rolling average) as candidate boundaries between LD blocks. This method prioritizes genomic regions with consistently low LD as natural split points while enforcing constraints on minimum and maximum block size. The resulting LD sets provide a flexible and data-driven foundation for modular fine-mapping across the genome, enhancing both resolution and computational tractability.

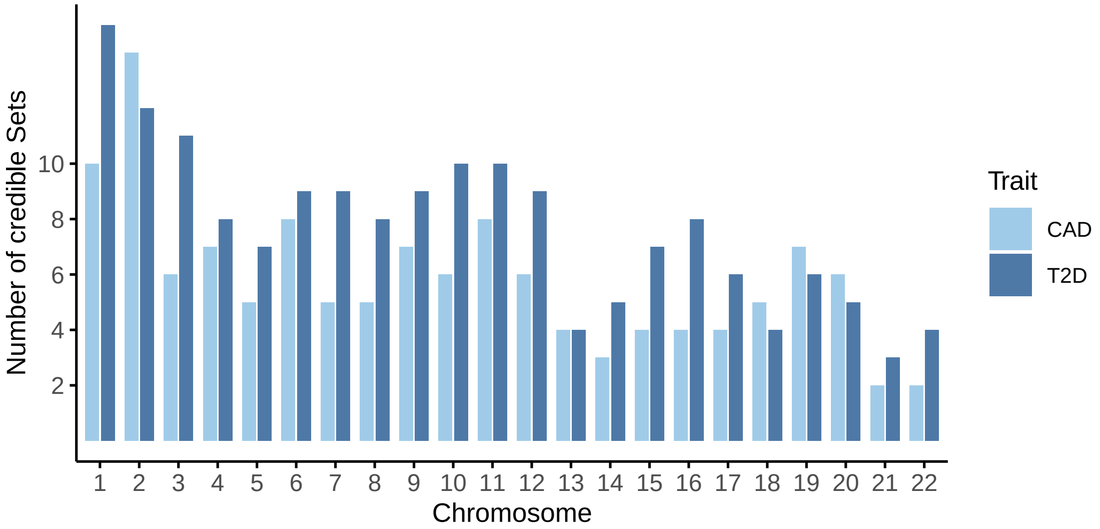

**Figure S5: Number of fine-mapped credible sets per chromosome for T2D and CAD.** Bar plot showing the number of distinct credible sets identified through Bayesian fine-mapping across autosomal chromosomes for type 2 diabetes (T2D, dark blue) and coronary artery disease (CAD, light blue). Each bar represents the count of credible sets on a given chromosome, with colors distinguishing the two traits.

We identified 169 credible sets for T2D and 128 credible sets for CAD (see Supplementary Tables SX and SXX), each representing a genomic region with a high probability of containing one or more causal variants. These credible sets provide a more precise localization of genetic signals beyond traditional GWAS loci, and facilitate downstream interpretation and functional characterization. The distribution of credible sets across chromosomes is summarized in Figure S5, highlighting both shared and trait-specific patterns of genetic architecture.

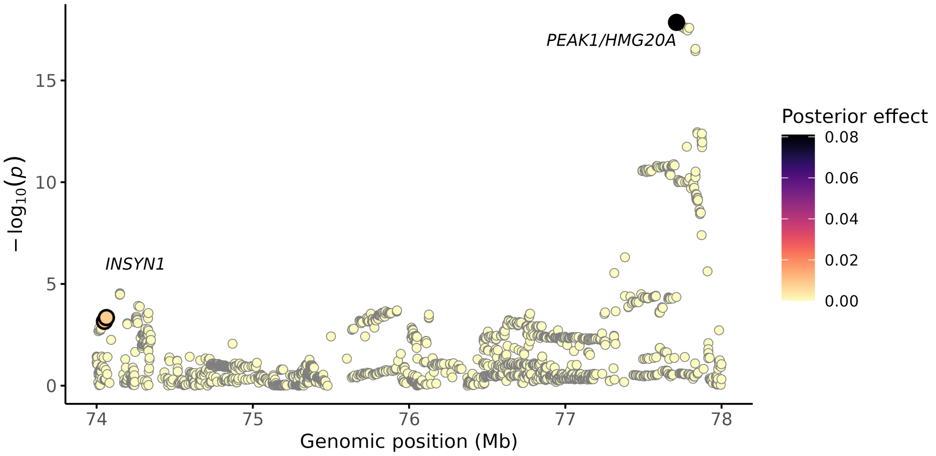

**Figure S6: Fine-mapping locus plot of chromosome 15 for Type 2 Diabetes (T2D)**. This plot displays association signals across the chromosome 15 locus, highlighting single nucleotide polymorphisms (SNPs) with their posterior effect sizes (colored and sized by Bayesian marker effect estimates). The x-axis represents genomic position (Mb), and the y-axis shows -log10 $P$-values from GWAS summary statistics. Credible set SNPs with high posterior inclusion probabilities are emphasized to pinpoint likely causal variants. Key genes within these two loci, potentially implicated in T2D pathogenesis, are annotated on the plot.

As an illustrative example of our fine-mapping results, Figure S6 displays the locus plot for chromosome 15 in the T2D analysis, highlighting two distinct credible sets. These sets localize association signals to genomic regions containing the gene *INSYN1* in one set, and *PEAK1* and *HMG20A* in the other CS.

Among the fine mapped loci we found 3 unique SNPs to be shared among T2D and CAD. In addition, among all the identified CS, 84 genes were tagged by a CS for both T2D and CAD.

### 7 Polygenic scoring

#### 7.1 Global and pathway-specific PGS

Polygenic scores (PGS) quantify an individual's genetic propensity for a given trait or disease by aggregating the effects of many genetic variants into a single predictive measure. As complex traits such as type 2 diabetes (T2D) and coronary artery disease (CAD) are influenced by thousands of small-effect variants, PGS have become an essential tool for risk prediction, genetic stratification, and personalized medicine.

Traditional PGS methods treat all associated variants equally, regardless of their biological context. However, recent advances suggest that biologically informed or pathway-partitioned PGS can provide additional insights into the underlying aetiology of disease, enhance interpretability, and potentially improve predictive performance. Partitioned PGS allow us to investigate how specific molecular pathways or regulatory annotations contribute to overall genetic risk, helping to dissect the polygenic architecture of complex traits.

Box 7.1: Statistical Models for Polygenic Score Construction

**Clumping and Thresholding (C+T)**

The classic PGS approach is based on a two-step clumping and thresholding procedure:

- Clumping: Select the most significant variant in a region and remove nearby variants in high linkage disequilibrium (LD), typically using a predefined $r^2$ threshold and window size.
- Thresholding: Define a $P$-value threshold $P_T$ below which SNPs are included in the score.

The polygenic score for individual $i$ is computed as:

$$\mathrm{PGS}_i = \sum_{j=1}^{m} \hat{\beta}_j \cdot G_{ij} \tag{21}$$

Where:

- $G_{ij}$ is the genotype of individual i at SNP $j$,
- $\hat{\beta}_j$ is the GWAS-derived effect size estimate,
- and $m$ is the number of included SNPs after clumping and thresholding.

While straightforward and computationally efficient, C+T methods do not fully account for LD or estimation uncertainty and may underperform in highly polygenic settings.

**Bayesian Shrinkage Models**

Bayesian models offer a more principled framework by explicitly modeling uncertainty in SNP effects and the LD structure. In this context, we focus on BayesC and BayesR priors, both implemented in our ‘gact‘ package for PGS construction.

**BayesC** assumes that only a small proportion of SNPs have non-zero effects. This is modelled via a spike-and-slab prior:

$$\beta_j \sim \begin{cases} 0 & \text{with probability } 1-\pi \\ \mathcal{N}(0, \sigma_\beta^2) & \text{with probability } \pi \end{cases} \tag{22}$$

Where:

- $\beta_j$ is the true SNP effect,
- $\pi$ controls sparsity (typically small, e.g., 0.01),
- $\sigma_\beta^2$ is the variance of non-zero effects.

This model allows joint estimation of SNP effects and borrowing of information across markers via LD.

**BayesR** extends BayesC by modelling SNP effects as a mixture of multiple normal distributions:

$$\beta_j \sim \sum_{k=1}^{K} \pi_k \cdot \mathcal{N}(0, \sigma_k^2) \tag{23}$$

Where:

- $\pi_k$ are the mixture proportions (e.g., for null, small, medium, and large effects),
- $\sigma_k^2$ are the corresponding variances,
- $K$ typically equals 4, with one component fixed at $\sigma^2 = 0$ for null effects.

This richer prior allows more flexible modelling of the heterogeneity in effect sizes, improving both estimation accuracy and predictive performance in polygenic traits.

Bayesian methods yield posterior mean effect estimates, which can be used directly to construct PGS:

$$\text{PGS}_i = \sum j = 1^m \mathbb{E}[\beta_j] \cdot G_{ij} \tag{24}$$

**Pathway-Partitioned PGS**

Using the gene set annotations available within gact, we can partition the genome into biologically meaningful subsets, such as Reactome pathways, tissue-specific gene sets, or regulatory features. For each subset, a pathway-specific PGS is computed:

$$\text{PGS}_i^{(k)} = \sum j \in S_k \mathbb{E}[\beta_j] \cdot G_{ij} \tag{25}$$

Where:

- $S_k$ is the set of SNPs linked to pathway $k$,
- $\text{PGS}_i^{(k)}$ represents the contribution from pathway $k$ to the individual's total genetic liability.

This approach enables the dissection of genetic risk by biological function, facilitating both mechanistic understanding and trait subtyping.

##### 7.2 Polygenic prediction of T2D and CAD

We computed polygenic scores (PGS) for T2D and CAD using the Bayesian linear regression (BLR) output from fine-mapping. Fine-mapping - and, consequently, the accuracy of polygenic prediction - is sensitive to misspecified linkage disequilibrium (LD). This is particularly relevant when using GWAS summary statistics from meta-analyses, where LD mismatches between reference panels and summary data may occur. We, and others (Shrestha et al., 2025; Wu et al., 2025), have proposed using the MCMC eigenvalue decomposition to address these discrepancies.

Importantly, the precision of LD estimation also depends on minor allele frequency (MAF). To mitigate LD misspecification, we performed BLR-based fine-mapping within predefined LD sets, using our built-in function ´createLDsets'. This function partitions genetic variants into approximately independent LD blocks based on smoothed LD scores. It identifies regions of low LD via a moving average and defines genome-wide split points while avoiding overly close cuts. These LD blocks are well suited for downstream applications such as fine-mapping, PGS construction, and enrichment testing, where LD independence is crucial for statistical validity.

For comparison, we evaluated prediction performance using both our custom-defined LD sets and those generated by the algorithm from Berisa and Pickrell (Berisa & Pickrell, 2015). Their method infers approximately independent LD blocks by analysing the patterns of correlation between genetic variants ($r^2$) in population-scale reference panels. Specifically, the genome is partitioned by identifying local minima in a smoothed $r^2$ correlation matrix, effectively pinpointing recombination cold spots that define block boundaries.

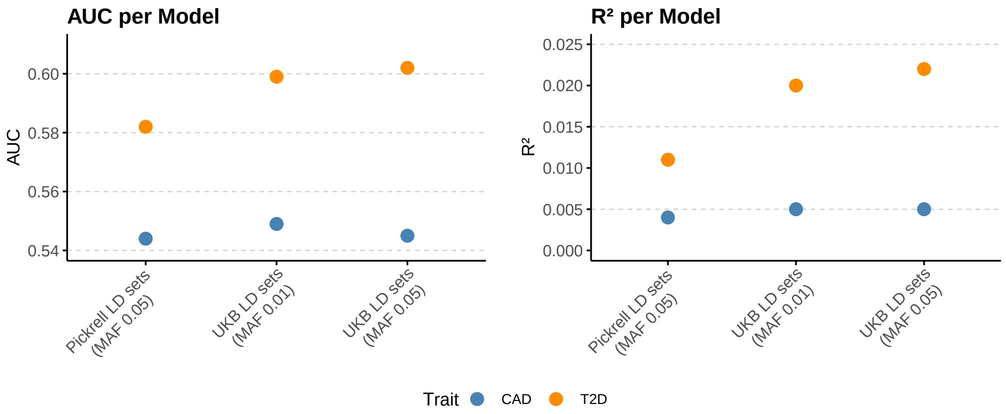

**Figure S7: Predictive performance of polygenic scores (PGS) for coronary artery disease (CAD) and type 2 diabetes (T2D)**. The accuracies are summarised across different linkage disequilibrium (LD) reference panels and minor allele frequency (MAF) thresholds. Left panel shows area under the curve (AUC); right panel shows Nagelkerke's $R^2$. Points represent model performance using UK Biobank (UKB) or Pickrell LD reference sets, with MAF thresholds of 0.01 or 0.05. Colors indicate the trait predicted: CAD (blue) and T2D (orange).

From Figure S7, it is evident that both the definition of LD sets and the MAF cut-off used for selecting genetic variants to estimate LD strongly influence the subsequent accuracy of polygenic prediction.

We further explored the genetic architecture of T2D by constructing pathway-partitioned PGS using Reactome-defined pathways. This approach allows us to evaluate whether specific biological processes disproportionately contribute to disease risk. A total of 33 pathways showed significant enriched PGS in T2D patients compared with controls S8, highlighting distinct molecular mechanisms associated with disease susceptibility. By integrating pathway-level information, our framework enables biologically interpretable PGS analyses that can inform downstream functional studies and enrichment tests.

#### 8 Maintaining and updating the `gact` package

The `gact` package has been designed with modularity and flexibility to support continuous integration of new data resources and methodological advances. Below, we outline recommended practices for maintaining and updating the package. The package has two key features:

1. Preprocessing of GWAS summary statistics, and

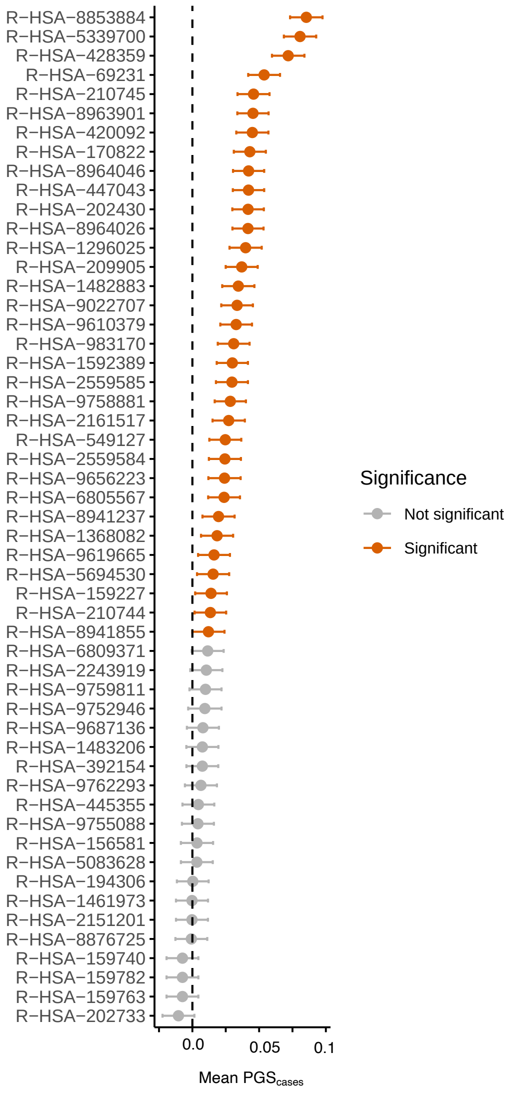

**Figure S8: Pathway partitioned polygenic scores (PGS) for type 2 diabetes (T2D)**. Enrichment of genetic burden across top 20 Reactome pathways in T2D patients compared to controls; significantly enriched pathways are highlighted in orange.

2. Linking common genetic variation to a range of biological annotations and features (e.g., genes, gene sets, pathways, and drug targets).

In the earlier sections, we demonstrated how the database can be updated with new GWAS summary statistics. For example:

```
GAlist <- updateStatDB(GAlist = GAlist,
                       stat = stat,
                       source = "Mahajan.NatGenet2018b.T2D-noUKBB.European.zip",
                       trait = "T2D",
                       type = "binary",
                       gender = "both",
                       ancestry = "EUR",
                       build = "GRCh37",
                       reference = "PMID:30297969",
                       n = 456236,
                       ncase = 55927,
                       ncontrol = 400309,
                       comments = "Exclude UK biobank",
                       writeStatDB = TRUE)
```

Similarly, existing GWAS summary data can be removed from the database using:

When the database is initialized for the first time, not all available gene sets are necessarily included. To add or update gene set annotations in the database, users can call the `createMarkerSetsDB()` function. The what argument can be used to specify which specific annotations should be added or refreshed:

```
GAlist <- createMarkerSetsDB(GAlist = GAlist, what="reactome")
```

#### Citered literature

Berisa, T., & Pickrell, J. K. (2015). Approximately independent linkage disequilibrium blocks in human populations. *Bioinformatics*, *32*(2), 283–285. https://doi.org/10.1093/bioinformatics/btv546

Bulik-Sullivan, B. K., Loh, P.-R., Finucane, H. K., Ripke, S., Yang, J., Patterson, N., Daly, M. J., Price, A. L., & Neale, B. M. (2015). LD Score regression distinguishes confounding from polygenicity in genome-wide association studies. *Nature Genetics*, *47*(3), 291–295. https://doi.org/10.1038/ng.3211

Cui, R., Elzur, R. A., Kanai, M., Ulirsch, J. C., Weissbrod, O., Daly, M. J., Neale, B. M., Fan, Z., & Finucane, H. K. (2024). Improving fine-mapping by modeling infinitesimal effects. *Nature Genetics*, *56*(1), 162–169.

Gholipourshahraki, T., Bai, Z., Shrestha, M., Hjelholt, A., Hu, S., Kjolby, M., Rohde, P., & Sørensen, P. (2024). Evaluation of Bayesian Linear Regression models for gene set prioritization in complex diseases. *PLOS Genetics*, *20*(11), e1011463. https://doi.org/10.1371/journal.pgen.1011463

Liu, J., McRae, A., Nyholt, D., Medland, S., Wray, N., Brown, K., Hayward, N., Montgomery, G., Visscher, P., Martin, N., & Macgregor, S. (2010). A versatile gene-based test for genome-wide association studies. *American Journal of Human Genetics*, *87*(1), 139–145.

Mahajan, A., Taliun, D., Thurner, M., Robertson, N. R., Torres, J. M., Rayner, N. W., Payne, A. J., Steinthorsdottir, V., Scott, R. A., Grarup, N., Cook, J. P., Schmidt, E. M., Wuttke, M., Sarnowski, C., Mägi, R., Nano, J., Gieger, C., Trompet, S., Lecoeur, C., … McCarthy, M. I. (2018). Fine-mapping type 2 diabetes loci to single-variant resolution using high-density imputation and islet-specific epigenome maps. *Nature Genetics*, *50*(11), 1505–1513. https://doi.org/10.1038/s41588-018-0241-6

Mishra, A., & Macgregor, S. (2015). VEGAS2: Software for more flexible gene-based testing. *Twin Research and Human Genetics*, *18*(1), 86–91.

Rohde, P. D., Sørensen, I. F., & Sørensen, P. (2019). qgg: an R package for large-scale quantitative genetic analyses. *Bioinformatics*, *36*(8), 2614–2615. https://doi.org/10.1093/bioinformatics/btz955

Rohde, P. D., Sørensen, I. F., & Sørensen, P. (2023). Expanded utility of the R package, qgg, with applications within genomic medicine. *Bioinformatics*, *39*(11), btad656. https://doi.org/10.1093/bioinformatics/btad656

Shrestha, M., Bai, Z., Gholipourshahraki, T., Hjelholt, A. J., Kjølby, M., Rohde, P. D., & Sørensen, P. (2025). Enhanced genetic fine mapping accuracy with Bayesian linear regression models in diverse genetic architectures. *PLOS Genetics*, *7*(21), e1011783. https://doi.org/10.1371/journal.pgen.1011783

Wu, Y., Zheng, Z., Thibaut, L., Lin, T., Feng, Q., Cheng, H., Yengo, L., Goddard, M. E., Wray, N. R., Visscher, P. M., & Zeng, J. (2025). Genome-wide fine-mapping improves identification of causal variants. *medRxiv*. https://doi.org/10.1101/2024.07.18.24310667
